# RNA Splicing of the REST Gene by SRRM4 in Vascular Smooth Muscle Cells Promotes Abdominal Aortic Aneurysm Development

**DOI:** 10.64898/2026.09.21.26363614

**Authors:** Yinan Li, Yuli Wang, Qihong Ni, Shuofei Yang, Haozhe Qi, Yongjie Yao, Hai Wang, Wei Xu, Kai Yuan, Lei Lv, Xiangjiang Guo, Meng Ye, Guanhua Xue, Wai Ho Tang, Jun Pu, Lan Zhang

## Abstract

**Background:** The progression of abdominal aortic aneurysm (AAA) is closely associated with the phenotypic switch of vascular smooth muscle cells (VSMC). While members of the Serine/Arginine-rich splicing factor (SRs) superfamily are increasingly acknowledged for their pivotal role in modulating cell phenotype, the specific contribution of these factors to AAA development has remained underexplored.

**Methods:** Through integrated analyses the whole transcriptome of human and murine AAA tissues derived from clinical samples and animal models, we identified serine-arginine repeat matrix 4 (SRRM4) as a candidate gene regulating AAA formation. To investigate the role of SRRM4 in AAA formation, we used global and VSMC-specific SRRM4-knockout mice to construct AAA mouse models in two different ways. The impact of SRRM4 on AAA was further investigated in *in vivo* and in *vitro* models

**Results:** Elevated levels of SRRM4 mRNA and protein were observed in VSMC from both human and murine AAA tissues. Targeted silencing of SRRM4 in VSMC (but not in endothelial or myeloid cells) significantly attenuated AAA formation in both AngII- and CaCl_2-_induced mouse models. Our results identify the RE1 silencing transcription factor (REST) as a critical target gene of SRRM4 in AAA, a master regulator of cellular phenotypic stability. We show that SRRM4 modulates VSMC phenotype through alternative splicing of the REST gene, resulting in a non-functional REST4 isoform. Furthermore, our study shows that REST directly represses STAT1, a mechanism by which SRRM4 influences VSMC phenotypic transition. Regarding clinical significance, inhibition of both STAT1 expression and function was shown to be beneficial in preventing AAA formation and slowing disease progression in mouse models.

**Conclusions:** This investigation uncovers a previously unrecognized role of SRRM4-mediated splicing of the REST gene in the progression of AAA and identifies a novel REST/STAT1 axis critical in the phenotypic modulation of VSMC. The findings of this study may provide insights for the development of innovative therapeutic strategies for AAA.

**What is New?:**

1. Our study demonstrates that the RNA splicing factor SRRM4 (serine/arginine repetitive matrix 4) expression is increased in human and murine abdominal aortic aneurysm (AAA) tissues and unveils a previously undescribed role of SRRM4 in regulating AAA development.
2. SRRM4 promotes AAA development and progression through inducing splicing of REST (RE1 silencing transcription factor) gene and upregulating STAT1 (signal transducer and activator of transcription 1) expression, which result in extracellular matrix degradation, inflammation, and VSMC phenotypic switching.
3. Our study provides proof-of-concept evidence indicating the therapeutic value of STAT1 inhibition in AAA prevention and treatment.

**What Are the Clinical Implications?:**

1. Our study defining a novel mechanism underlying the SRRM4/REST/STAT1 axis is crucial for developing new therapeutic strategies for AAA.
2. Based on these findings, we have initiated a prospective study (REST4-AAA; ClinicalTrials.gov Identifier: NCT07056491) to evaluate REST4 expression levels in serum of AAA patients and explore its potential as a serum biomarker for AAA prediction
3. Our findings indicate that fludarabine, a clinically available agent reported to selectively suppress STAT1, effectively attenuates AAA progression in mice, suggesting its potential for repurposing as a therapeutic strategy for AAA.

## INTRODUCTION

Aortic aneurysm is a matrix degenerative disease characterized by the dilatation of a blood vessel. Such aneurysms compromise the integrity of the arterial wall, increasing the risk of rupture and resulting in extensive and often fatal internal bleeding^1, 2^. The vast majority of AAAs remain asymptomatic until they progress rapidly to rupture, making early diagnosis difficult. Although endovascular stent grafting is widely used for AAA treatment, no pharmacological interventions are currently capable of halting AAA progression or reducing rupture risk.^1, 2^. Identifying novel therapeutic targets requires a comprehensive understanding of the molecular mechanisms governing AAA formation and progression, which is essential for advancing treatment modalities.

The sequential pathophysiology of aneurysm formation is unclear, but it is believed that vascular smooth muscle cells (VSMC) play a central role^3–5^. Most VSMC in the vessel wall display a contractile phenotype, which allows them to maintain vascular tone. However, VSMC have the ability to transdifferentiate into a synthetic phenotype. This process is termed phenotypic switching and is considered to be a key mechanism in vascular remodeling^4–6^. Synthetic VSMC are characterized by decreased contractile protein expression and increased the production of elastolytic enzymes such as matrix metalloproteinases (MMPs), which degrade the extracellular matrix (ECM) and facilitate migration by detaching cells from the basement membrane and ECM, all of which contribute to aortic wall weakness and AAA generation^7–9^. While our group has previously demonstrated that nuclear receptor-mediated transcriptional regulation in VSMC plays a pivotal role in AAA pathogenesis and progression^10–12^, it remains elusive whether alternative splicing participates in the modulation of VSMC phenotypic switching and subsequent AAA development.

RNA sequencing (RNA-seq) can elucidate molecular mechanisms of AAA development. Although recent work has focused on the genomic and transcriptomic profiles of AAA, analyses of alternative splicing have lagged. Through integrated analyses of RNA-seq results from human and murine AAA, we identified specific alternative splicing signatures of AAA. Although alternative splicing of several genes has been reported to correlate with AAA, none have been confirmed as driving AAA development and progression. The Serine/Arginine-rich (SR) splicing factor superfamily comprises splicing factors that orchestrate alternative splicing in various physiological and pathological states.^13–16^. Within this superfamily, several members have emerged as critical modulators of cell phenotype modulation. SRRM4 (also known as nSR100), a member of the SR superfamily, was initially identified as a key regulator of neurogenesis^16, 17^. Recent investigations have revealed novel functions of SRRM4 beyond its established roles in the neural system, including facilitation of neuroendocrine trans-differentiation and modulation of immune responses in prostate tumors^18–20^. Despite the observed expression of SRRM4 in non-neural tissues, its functional significance in the pathogenesis of AAA remains to be elucidated.

In our previous investigation, we elucidated the reduction of the RE1 silencing transcription factor (REST) in the synthetic phenotype of VSMC in atherosclerotic tissues^21^. REST is highly expressed in embryonic stem cells and non-neuronal cells and acts as a key negative regulator of neurogenesis by inhibiting genes essential for cellular differentiation^22, 23^. A notable strategy by which cells attenuate REST functionality involves alternative splicing of REST pre-RNA into REST4, where a specific exon (exon N) is inserted between exons 3 and 4, resulting in the translation of a truncated and functionally altered REST4 protein^19, 24^. These observations suggest that while transcriptomic analyses can pinpoint genes such as REST that are involved in VSMC phenotypic transition, the identification of RNA splicing factors associated with AAA may reveal previously unrecognized mechanisms underlying AAA progression and potentially innovative therapeutic strategies.

In the current study, we identify a role for SRRM4 in AAA pathogenesis and investigate the underlying mechanisms. SRRM4 facilitates VSMC phenotypic transition by alternative splicing of REST into the functionless REST4 isoform. Loss of REST function leads to STAT1 upregulation, resulting in MMP2 activation and VSMC phenotypic switching. Targeting either SRRM4 or STAT1 prevented AAA progression and alleviated the overall pathological process.

## MATERIAL AND METHODS

### Human Tissue Collection

Human aortic tissues and plasma samples were collected from patients undergoing open AAA repair at Renji Hospital, Shanghai Jiao Tong University School of Medicine. Control aortic tissues were obtained from brain-dead organ donors, and age- and sex-matched healthy controls were enrolled for serum collection. All procedures were approved by the Research Ethics Committee of Renji Hospital (KY2020-151 and RA-2020-071), and written informed consent was obtained from all participants. Detailed clinical characteristics are provided in Table S1

### Animal Studies

All animal protocols were approved by the Ethics Committee of Renji Hospital (RJ2019-0320) and conducted in accordance with the National Institutes of Health Guidelines for the Care and Use of Laboratory Animals. VSMC-specific Srrm4 knockout mice, DOX-inducible VSMC-specific *Srrm4* knockdown mice, tamoxifen-inducible VSMC-specific *Srrm4* knockout mice, VSMC-specific *Stat1* knockout mice, and REST splicing mutant mice were generated as described in the Supplementary Methods. AAA was induced by AngII infusion or periaortic CaCl2 application. Detailed protocols are provided in the Supplementary Methods.

### Cell Studies and Molecular Assays

Primary mouse aortic VSMCs, endothelial cells, and macrophages were isolated and cultured as previously described. Human primary aortic smooth muscle cells were purchased from ATCC. Cell transfection, RNA sequencing, RNA immunoprecipitation, RNA pulldown, ChIP, dual-luciferase reporter assays, RT-qPCR, immunoblotting, immunohistochemistry, immunofluorescence, RNA in situ hybridization, and ELISA were performed as detailed in the Supplementary Methods.

### Statistical Analysis

All statistical analyses were performed using GraphPad Prism version 10.0 or SPSS version 20.0. The distribution of continuous variables and the homogeneity of variances were assessed before statistical testing. Because all comparisons involved independent samples, two-group comparisons were performed using a two-tailed, unpaired Student’s t test for normally distributed data. Welch’s t test was used when the assumption of equal variances was not met. For comparisons involving more than two independent groups, one-way analysis of variance (ANOVA) was used when a single factor was evaluated, whereas two-way ANOVA was used when two independent factors were examined. Post hoc comparisons following two-way ANOVA were performed with Bonferroni correction. Welch’s ANOVA was applied when the assumption of homogeneity of variances was violated. For data that did not meet the assumptions of parametric testing, the Mann-Whitney U test was used to compare two independent groups, and the Kruskal-Wallis test was used to compare more than two independent groups. Associations between the expression levels of two genes were assessed using Pearson’s correlation analysis. Bivariate associations between categorical variables were evaluated using the χ² test. Survival distributions were estimated using the Kaplan-Meier method and compared using the stratified log-rank test. Unless otherwise specified, normally distributed continuous variables are presented as the mean ± standard deviation (SD). All statistical tests were two-sided, and a p-value <0.05 was considered statistically significant.

### Data Sharing Statement

All data supporting the findings of this study are available from the corresponding author upon reasonable request.

## RESULTS

### SRRM4 is upregulated in VSMC of AAA tissues

We performed comprehensive transcriptome sequencing on aortic tissues from clinical AAA samples and an AngII-induced AAA animal model to investigate alternative splicing profiles. The analysis revealed distinct alternative splicing patterns in AAA tissues. In human samples, 106 out of 1036 predicted alternative splicing (AS) events from 916 genes exhibited statistically significant differences between AAA and non-AAA tissues (Benjamini-Hochberg corrected, FDR < 0.01). In the animal model, 1549 differentially expressed genes were identified between AAA and control samples, a finding that parallels the transcriptomic reprogramming previously observed in SRRM4-driven neuroendocrine prostate cancer versus prostate adenocarcinoma models. Among 1023 predicted AS events from 889 genes, 59 events showed significant differences (Benjamini-Hochberg corrected, FDR < 0.01) (Figure 1A & S1A). Among these AAA-specific splice variants, 16 genes have been reported to be regulated by the SR family of splicing factors, which includes SRSF1-11 and SRRM1-4. However, the role of most SRs in AAA remains unexplored. RNA-seq profiling of all SR genes in both clinical AAA samples and AngII-induced mouse models identified SRRM4 as the member with the highest fold change (Figure 1B). We then constructed a phenotype switch reporting system comprising two complementary fluorescent reporters: eGFP driven by the synthetic biomarker VIM and mCherry driven by the contractile biomarker α-SMA (Figure S2A). Screening of SR superfamily siRNAs in VSMCs carrying this reporter system showed that siRNA targeting SRRM4 produced the highest eGFP/mCherry repression rate (Figure S2B). This result suggests that SRRM4 plays an important role in AAA development and VSMC phenotype switching.

**Figure 1.**
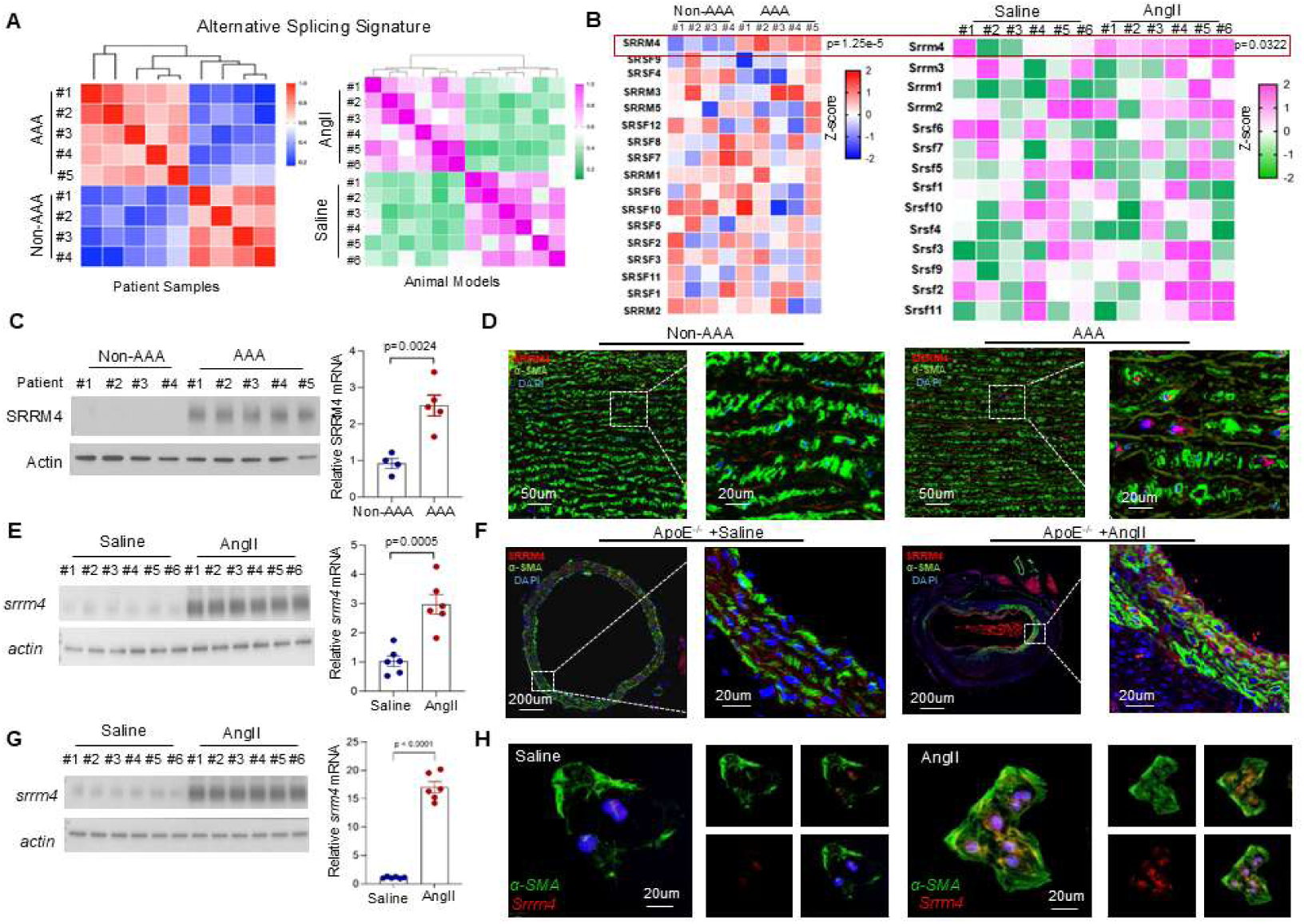
SRRM4 expression is upregulated in VSMC of human and mice AAA tissues. **A)** Heat maps showed Spearman correlation values using the splicing index of samples from the human AAA and non-AAA segments and mouse abdominal aortic segments. Clustering is performed as in panel, based on the entire set of splicing predictions. **B)** Relative mRNA expression levels of SR superfamily members were measured by RNA-seq profiling in control of AAA tissues (left) and saline- or AngII-induced AAA mice (right). The raw data were calculated and presented as Z-score. **C)** Quantification of SRRM4 mRNA and protein expression measured by realtime-qPCR and immunoblotting in human AAA and non-AAA segments. **D)** Representative images of SRRM4 protein expression by IF staining of human AAA and non-AAA segments as described in the M&M section. **E)** Quantification of *Srrm4* mRNA and protein expression measured by realtime-qPCR and immunoblotting in mouse abdominal aortic segments. **F)** Representative images of *Srrm4* protein expression by IF staining of mouse abdominal aortic segments as described in the M&M section. **G)** Quantification of *Srrm4* mRNA and protein expression measured by realtime-qPCR and immunoblotting in primary cultured VSMC derived from mouse abdominal aortic segments after stimulation with PBS or AngII for 48 hours. **H)** Representative images of *Srrm4* expression by immunofluorescence staining in primary cultured VSMC derived from mouse abdominal aortic segments and containing with α-SMA and DAPI.

To investigate the expression profile of SRRM4 in AAA, we first used an AngII-induced AAA mouse model. Immunoblotting and RT-qPCR revealed significantly increased SRRM4 abundance in both human and murine AAA tissues (Figure 1C & 1E). RT-qPCR of plasma exosomes from 154 AAA patients and 234 non-AAA controls showed significantly increased SRRM4 mRNA levels in AAA patients (Figure S2C). Immunofluorescence staining of aortic sections revealed markedly increased SRRM4 protein, predominantly in smooth muscle cells, in both human and murine AAA tissues (Figure 1D & 1F). Increased alternative splicing of SRRM4-target genes (ABI1, PTK2) was also observed in both species (Figure S3). In primary cultured VSMCs from mouse AAA tissues, SRRM4 mRNA and protein levels were significantly increased upon AngII treatment (Figure 1G-H). SRRM4 expression was upregulated as early as 3 days in AngII-induced aortic tissues (Figure S4A), whereas no change was observed in primary cultured endothelial cells or macrophages (Figure S4B). These results suggest a potential role for SRRM4 in AAA pathology.

To investigate whether this upregulation could be attributed to common genetic predisposition, we analyzed the association between SNPs at the SRRM4 locus and AAA risk using large-scale GWAS data. No common SNP within the SRRM4 gene region showed significant association with AAA susceptibility, indicating that the observed SRRM4 dysregulation is unlikely to be driven by inherited genetic variants (Figure S5A). We further investigated the upstream signaling pathways leading to SRRM4 activation in AAA and found that promoter unmethylation may contribute to SRRM4 upregulation in AAA pathogenesis (Figure S5B-E). Taken together, our data indicate that SRRM4 is upregulated in aortic VSMCs in AAA tissue.

### VSMC-specific silencing of SRRM4 represses AAA formation

Upregulation of SRRM4 was only observed in VSMCs (Figure 1 & S4), so we next focused on the role of VSMC-specific SRRM4 in AAA formation. To determine whether SRRM4 contributed to AAA progression, we constructed the VSMC-specific Srrm4 knockout *ApoE^−/−^* mice *(ApoE*^−/−^/*Srrm4^ΔSMC^*) and subjected them to AngII-induced AAA (Figure 2A and S6A-B). Under non-treatment conditions, SMC-specific Srrm4 silencing did not significantly alter the physical properties of the aorta, with no significant differences in HE staining, elastin degradation, aortic blood flow, KCl-induced contraction, or acetylcholine-induced relaxation between wild-type and Srrm4-deficient mice (Figure S6C-G). However, upon AngII stimulation, the *ApoE*^−/−^/*Srrm4^ΔSMC^* mice were less sensitive to AngII induction and had smaller aneurysms, with significant reductions in mortality rate, AAA incidence ratio, and mean maximum diameter compared with controls (Figure 2B-G). Excess collagen deposition and elastin breakage were rescued by *Srrm4* silencing (Figure 2H). No significant difference was observed in systolic/diastolic blood pressure, heart rate and plasma lipids between *ApoE^−/−^*/*Srrm4^ΔSMC^* and *ApoE^−/−^*/*Srrm4^flox/flox^* mice (Figure S7). We confirmed these findings in a second CaCl2-induced AAA model, in which VSMC-specific *Srrm4* deletion prevented AAA development and inhibited medial elastic fiber fragmentation and aortic wall thinning or rupture (Figure S8). To further investigate which cell type contributed to the observed phenotype, we generated endothelial and myeloid cell-specific *Srrm4*-knockout mice (*ApoE*^−/−^/*Srrm4*^ΔEC^, *ApoE*^−/−^/*Srrm4*^ΔMΦ^), and their littermate controls (Figure S6B). However, phenotypes were minimally affected in these mice (Figure S9). These results indicate that VSMC-derived SRRM4 acts as a positive regulator in AAA pathogenesis.

**Figure 2.**
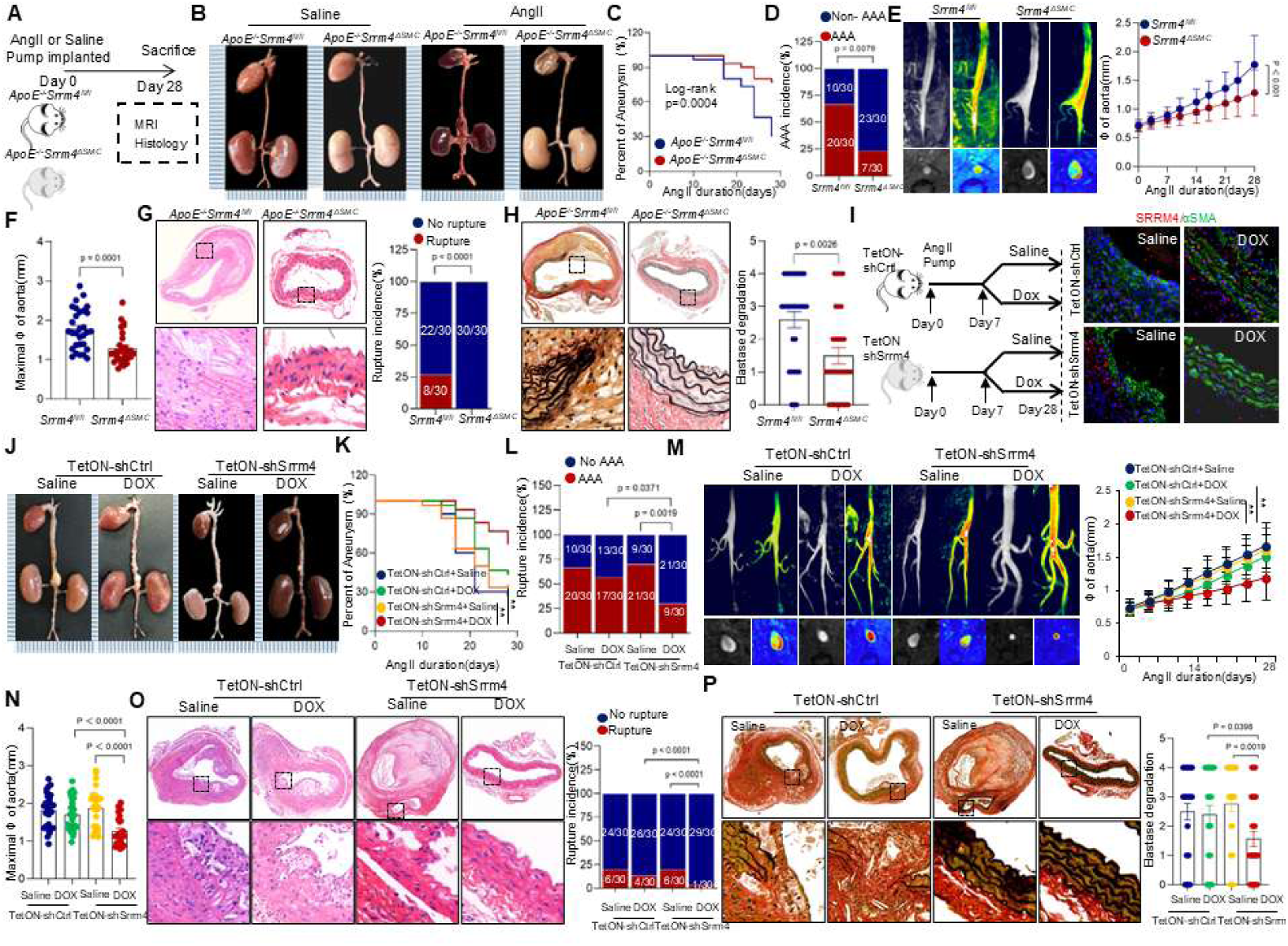
VSMC-specific silencing of SRRM4 represses AngII-induced AAA formation. **A)** Schematic protocol of: *ApoE^−/−^*/*Srrm4^flox/flox^* and *ApoE^−/−^*/*Srrm4^ΔSMC^* mice were subcutaneously injected with saline or AngII by a mini osmotic pump for 28 days. **B)** Representative images of the macroscopic features of AAA formation in indicated groups. **C)** Survival curves in indicated groups. Survival data were analyzed by the Kaplan-Meier method and compared using log-rank tests. **D)** The incidence of AngII-induced aneurysm in indicated groups were analyzed by the Fisher exact test. **E)** Representative images of abdominal aortas visualized by MRI-scan in indicated groups. Growth curve of diameter of suprarenal abdominal aortas measured by MRI-scan in indicated groups. **F)** Quantification of the maximal diameter of suprarenal abdominal aortas from indicated groups by MRI-scan. **G-H)** Representative H&E and EVG staining images of suprarenal abdominal aortas in indicated groups after 28 days of saline or AngII infusion. Quantification of elastin degradation by counting the number of breaks per vessel in suprarenal aortic segments, based on EVG staining. The rupture incidence of AngII-induced aneurysm in indicated groups were analyzed by the Fisher exact test. **I)** The transgenic mice with smooth muscle cell-specific knockdown of the SRRM4 protein induced by DOX were constructed as described in the Material and Methods. Schematic protocol of: AngII pumps were implanted subcutaneously in the mice, and after 7 days, they were fed either Saline or DOX. After 28 days, aortic tissue samples were collected. Immunofluorescence was used to verify the specific expression of the SRRM4 protein in smooth muscle cells. **J)** Representative images of the macroscopic features of AAA formation in indicated groups. **K)** Survival curves in indicated groups. Survival data were analyzed by the Kaplan-Meier method and compared using log-rank tests. **L)** The incidence of AngII-induced aneurysm in indicated groups were analyzed by the Fisher exact test. **M)** Representative MRI images of the abdominal aorta in the indicated groups. Growth curves showing the diameter of the suprarenal abdominal aorta measured by MRI in the indicated groups. **N)** Quantification of the maximal diameter of suprarenal abdominal aortas from indicated groups. **O-P)** Representative H&E and EVG staining images of suprarenal abdominal aortas in indicated groups after 28 days of saline or AngII infusion. Quantification of elastin degradation by counting the number of breaks per vessel in suprarenal aortic segments, based on EVG staining. The rupture incidence of AngII-induced aneurysm in indicated groups were analyzed by the Fisher exact test.

To validate the critical role of targeting SRRM4 in VSMCs, we generated a mouse model with DOX-inducible VSMC-specific Srrm4 silencing (*ApoE^-/-^*Tagln-rtTA; TRE3G-shRNA(Srrm4)) and subjected them to AngII-induced AAA (Figure 2I). Since previous studies reported a protective effect of DOX on AAA, we included additional control groups to rule out any effects of the inducing agent itself. DOX-induced VSMC-specific *Srrm4* silencing significantly ameliorated AAA formation and progression (Figure 2I-N), with reduced arterial wall thickening and elastin degradation, as well as decreased maximal aortic diameter and AAA rupture rate compared with saline-treated mice (Figure 2O-P).

To further validate that the observed protection is VSMC-intrinsic and independent of the induction system used, we additionally generated tamoxifen-inducible VSMC-specific Srrm4 knockout mice (*ApoE^-/-^*;*Srrm4^flox/flox^*;*Tagln-CreERT2*). Mice were treated with tamoxifen followed by a 14-day washout period before AngII infusion to exclude any direct pharmacological effect of tamoxifen. Consistent with the DOX-inducible model, tamoxifen-induced Srrm4 knockout significantly attenuated AngII-induced AAA formation, with reduced AAA incidence, decreased maximal aortic diameter, and preserved elastin integrity (Figure S10). These findings confirm that SRRM4 functions cell-autonomously in VSMCs to promote AAA pathogenesis, independent of the induction system or inducing agent used. Taken together, our results demonstrate that VSMC-specific targeting of SRRM4 effectively delays AAA development and progression.

### SRRM4 promotes VSMC phenotype switch and ECM degradation

VSMC phenotypic switching is a key hallmark of AAA, characterized by a transition from a quiescent contractile phenotype to a proliferative synthetic state that triggers ECM remodeling and promotes a proinflammatory environment. We investigated the correlation between SRRM4 expression and the degree of VSMC phenotype switching and MMP expression in both human and murine AAA samples.

In human AAA samples, SRRM4 expression was positively correlated with the synthetic markers VIM and MMP2, and negatively correlated with the contractile marker α-SMA (Figure 3A-B & S11A). Consistent with this, SRRM4 knockdown in murine models decreased VIM and MMP2 levels and MMP2 activity, while increasing α-SMA expression in aortic tissues (Figure 3C-H & S11B-C). To corroborate the pro-synthetic role of SRRM4, we performed gain-of-function experiments by stably overexpressing SRRM4 in VSMCs. SRRM4 overexpression significantly upregulated the mRNA and protein levels of synthetic markers VIM, MMP2, and COL1A1, while downregulating the contractile markers α-SMA and Tagln (Figure 3I). Functionally, SRRM4 overexpression impaired VSMC contractile ability, as assessed by collagen gel contraction assay (Figure 3J), and markedly promoted MMP2 secretion (Figure 3K) and MMP activity (Figure 3L). These data demonstrate that SRRM4 is sufficient to drive VSMC phenotype switching and MMP2 activation.

**Figure 3.**
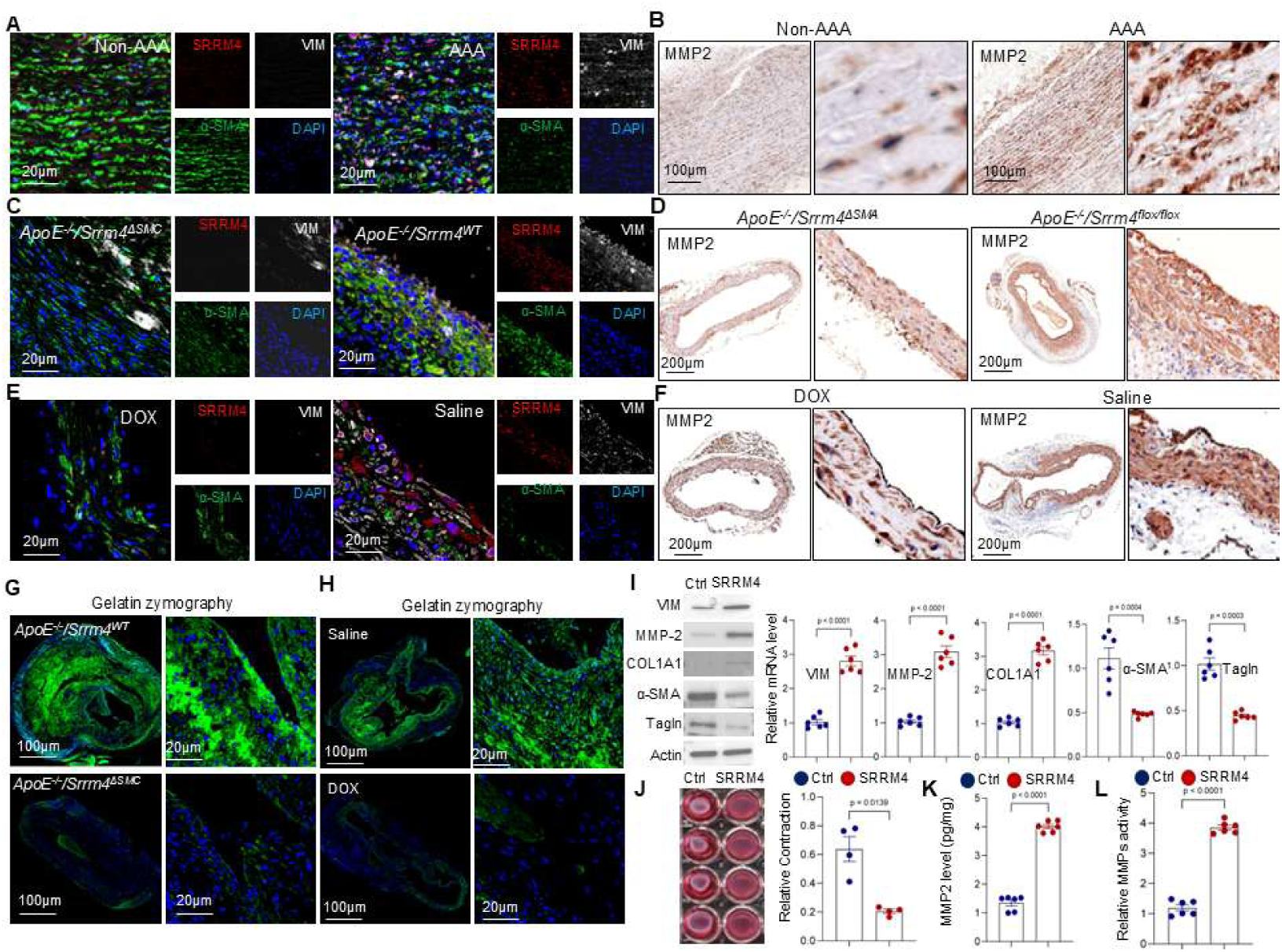
SRRM4 drives VSMC phenotype switch in AAA. **A-B)** A) IF staining using SRRM4, α-SMA, and VIM antibodies and B) IHC staining using MMP2 antibody were performed on human AAA and non-AAA segments, representative images of IF staining are shown. **C-D)** C) IF staining using SRRM4, α-SMA, and VIM antibodies and D) IHC staining using MMP2 antibody were performed on suprarenal abdominal aortas derived from *ApoE^−/−^*/*Srrm4^flox/flox^* and *ApoE^−/−^*/*Srrm4^ΔSMC^* mice, representative images of IF staining are shown. **E-F)** E) IF staining using SRRM4, α-SMA, and VIM antibodies and F) IHC staining using MMP2 antibody were performed on suprarenal abdominal aortas derived from Tagln-rtTA; TRE3G-shRNA(Srrm4) mice treated with saline or DOX, representative images of IF staining are shown. **G-H)** In-situ zymography (DQgelatin) was performed on aortas from G) *ApoE^−/−^*/*Srrm4^flox/flox^* and *ApoE^−/−^*/*Srrm4^ΔSMC^* and H) Tagln-rtTA; TRE3G-shRNA(Srrm4) mice treated with saline or DOX after AngII infusion as described in the Material and Methods section. Representative images are shown. **I-L)** VSMC were stably over expressed with control or SRRM4; I) Quantification of mRNA and protein levels of contractile markers α-SMA and Tagln, and the synthetic markers VIM, COL1A1, and MMP2 in VSMC and VSMC with SRRM4 overexpression; J) VSMC contraction ability were measured by collagen gel contraction assay; K) Secreted MMP2 level were measured by ELISA; L) MMP activity of indicated cells were measured by MMP activity assays.

To further substantiate the link between SRRM4 upregulation and VSMC phenotype switching, primary VSMCs isolated from aortic tissue of *ApoE^-/-^*/*Srrm4*^ΔSMC^ and *ApoE^-/-^*/*Srrm4*^flox/flox^ mice as well as VSMC-specific, DOX-inducible Srrm4 knockdown mice treated with saline or DOX were examined. Both immunoblotting and real-time quantitative PCR showed that VSMCs with SRRM4 silencing maintained a contractile phenotype and repressed MMP2 expression under AngII treatment (Figure S12A & E). SRRM4 silencing also significantly inhibited AngII-induced MMP2 secretion and activity (Figure S12B-D & F-H). Moreover, AngII augmented the levels of inflammatory molecules IL-18, IL-1β, and MCP-1 in aortic tissues from SRRM4-intact mice, whereas these levels were reduced in mice with SRRM4 silencing (Figure S13). SRRM4 knockdown reversed AngII-induced VSMC phenotype switching and inhibited AngII-induced MMP2 synthesis, secretion, and activation (Figure S14). Together, these results suggest that SRRM4 drives VSMC phenotype switching and induces matrix metalloproteinase and inflammatory factor synthesis, thereby promoting AAA development and progression.

### REST is the direct target of SRRM4 in regulating VSMC phenotype switch

To identify the target genes mediating SRRM4-induced VSMC phenotype switching, we performed RNA sequencing on VSMCs overexpressing SRRM4. Among genes with significantly altered alternative splicing events (FDR < 0.01), 23 of the 43 genes previously identified with altered splicing in AAA tissues^19^ also exhibited similar changes in SRRM4-overexpressing VSMCs (Figure 4A). Screening these 23 candidates using the phenotype switch reporter system revealed that antisense RNA interference with REST alternative splicing effectively blocked SRRM4-induced phenotype switching (Figure 4B). During REST splicing, exon N is inserted between exons 3 and 4, generating a truncated, functionally reprogrammed REST4 protein. SRRM4 overexpression significantly decreased REST while increasing REST4 protein and mRNA levels (Figure 4C). An antisense oligonucleotide (ASO) targeting the junction between intron 3 and exon N reversed SRRM4-induced phenotype switching (Figure 4D-H). Furthermore, transfection with an expression vector encoding a variant of the REST cDNA that is resistant to SRRM4-mediated splicing also attenuated the phenotype switch in VSMCs induced by SRRM4 (Figure S15A-D), while REST knockdown induced phenotype switch in VSMC (Figure S15E-H). These results indicate that SRRM4 drives VSMC phenotype switching largely through reprogramming REST function via alternative splicing.

**Figure 4.**
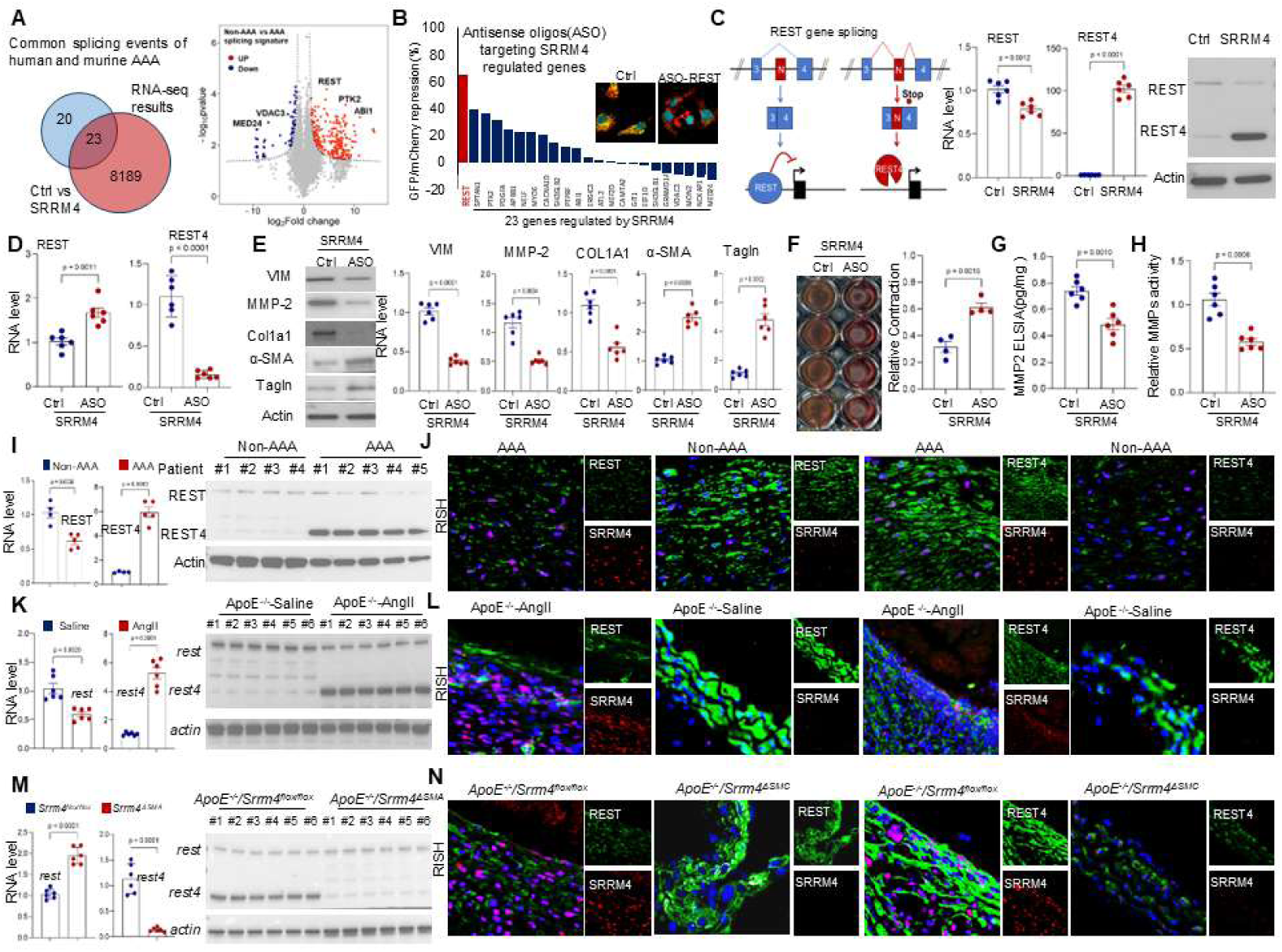
SRRM4 promotes VSMC phenotype switch through alternative splicing of REST gene. **A)** VSMCs overexpressing ctrl or flag-tagged SRRM4 were used to extract total RNA for RNA-seq analysis of genes with significantly altered alternative splicing. RNA-IP-seq was performed using flag antibodies to analyze SRRM4-binding genes, and a Venn diagram was used to show the 23 overlapped genes of the two groups. Total RNA derived from human AAA and non-AAA segments were used for RNA-seq, the alteration of gene splicing was showed as volcano plot based on log_2_ Fold change and -log_10_ p-value. **B)** SRRM4 and the VSMC phenotype switch reporting system were transfected into VSMCs, followed by the transfection of antisense oligonucleotides (ASOs) targeting splicing sites of the 23 genes to inhibit their alterative splicing. The suppression rate of GFP/mCherry fluorescence intensity was then measured. **C)** Diagram showed the alterative splicing of REST gene, expression of REST and REST4 in VSMC with or without SRRM4 expression were measured by immunoblotting and realtime-qPCR. **D-H)** VSMC stably overexpressed SRRM4 were treated with either control ASO or ASO targeting REST splicing; D) Inhibition of REST gene alteration splicing were validated by realtime-qPCR. E) Quantification of mRNA and protein levels of contractile markers α-SMA and TAGLN, and the synthetic markers VIM, COL1A1, and MMP2; F) VSMC contraction ability was measured by collagen gel contraction assay; G) Secreted MMP2 level was measured by ELISA; H) MMP activity of indicated cells was measured by MMP activity assays. **I)** Quantification of REST and REST4 expression were measured by realtime-qPCR and immunoblotting in human AAA and non-AAA segments. **J)** Representative images of SRRM4, REST, and REST4 mRNA expression by RISH staining of human AAA and non-AAA segments as described in the M&M section. **K)** Quantification of REST and REST4 expression were measured by realtime-qPCR and immunoblotting in abdominal aortic segments derived from *ApoE^−/−^* mice treated with saline or AngII. **L)** Representative images of *Srrm4*, *Rest* and *Rest4* mRNA expression by RISH staining of *ApoE^−/−^* mice treated with saline or AngII. **M)** Quantification of REST and REST4 expression were measured by realtime-qPCR and immunoblotting in suprarenal abdominal aortas of *ApoE^−/−^/Srrm4^flox/flox^* and *ApoE^−/−^/Srrm4^ΔSMC^* mice. **N)** Representative images of *Srrm4*, *Rest* and *Rest4* mRNA expression by RISH staining of *ApoE^−/−^/Srrm4^flox/flox^* and *ApoE^−/−^/Srrm4^ΔSMC^* mice as described in the M&M section.

We next examined REST and REST4 expression in human and mouse AAA models. Human AAA samples showed significantly reduced REST and increased REST4 levels compared to non-AAA counterparts (Figure 4I-J), with elevated REST4 mRNA also detected in patient blood exosomes (Figure S15I). This trend was paralleled in AngII-treated murine AAA models (Figure 4K-L). Notably, the REST4:REST ratio was significantly reduced in Srrm4-ablated mice compared to controls (Figure 4M-N and Figure S16A-B), highlighting a correlation between REST splicing and SRRM4 expression in AAA tissues. Together, these data identify REST as the direct target of SRRM4 in regulating VSMC phenotype switching.

We next explored how SRRM4 regulates REST splicing. RNA immunoprecipitation revealed that SRRM4 selectively recognizes the region proximal to the 3’ splice site of REST intron 3, with markedly reduced enrichment in the control intron region (Figure S16C). RNA pulldown confirmed direct interaction between SRRM4 and the UGC motif within REST intron 3 (Figure S16D). A REST minigene mimicked the native REST response to SRRM4 overexpression (Figure S16E), and site-directed mutagenesis showed that SRRM4-mediated exon N inclusion depends on guanine within this motif (Figure S16F). Thus, SRRM4 binding to the UGC motif facilitates exon N inclusion, leading to REST4 splicing.

To validate the role of REST splicing in AAA pathogenesis *in vivo*, we generated *ApoE^-/-^* mice with a G-A mutation in the SRRM4 binding site in intron 3 of the REST gene (*ApoE^-/-^* TAC-MT). Primary VSMCs were then isolated from these animals, and we found that the G-A mutation significantly disrupted REST gene splicing (Figure S17A-B). Compared to control VSMC(TGC-WT), VSMC(TAG-MT) maintained a contractile phenotype under AngII stimulation and exhibited reduced MMP2 synthesis and secretion (Figure S17C-F). In the AngII-induced AAA model (Figure S18A-B). The *ApoE^-/-^* VSMC-MT mice were less sensitive to AngII, with significant reductions in mortality, AAA incidence, and mean maximum diameter compared with controls (Figure S18C-F). Elastin breakage and VSMC phenotypic switching were also rescued by blocking REST splicing (Figure S18G-H). In summary, blocking SRRM4-induced REST alternative splicing suppressed VSMC phenotype switching and inhibited AAA progression.

### SRRM4/REST axis induced VSMC phenotype switch is mediated by STAT1

To elucidate the mechanisms by which REST splicing contributes to the pathogenesis of AAA, we analyzed the REST transcriptome in VSMC by ChIP-seq. Bioinformatic analysis of ChIP-seq data, coupled with public database exploration, identified 29 gene as potential downstream candidate of REST (Figure 5A). We further valid these genes using the VSMC phenotype switch reporting system and identified STAT1 as the crucial downstream target in REST splicing induced VSMC phenotype switch (Figure 5B-C). SRRM4 overexpression was found to increase STAT1 expression in VSMCs, while silencing of SRRM4 repressed STAT1 expression (Figure 5D). Immunobiochemical staining revealed increased STAT1 expression in human AAA tissues, where increased STAT1 protein levels were associated with increased VIM levels but decreased α-SMA protein levels (Figure 5E and S19A). This pattern was mirrored in AngII-treated AAA mouse aortas, which showed increased STAT1 and VIM in addition to decreased α-SMA (Figure 5F and S19A). Notably, SRRM4 knockdown resulted in decreased STAT1 expression in ApoE^-/-^/*Srrm4*^ΔSMC^ AAA mice compared to their counterparts (Figure 5G). These results indicate that STAT1 expression was governed by SRRM4/REST axis REST silencing increased STAT1 expression in VSMCs (Figure 5H). Both REST overexpression and ASO-mediated blockade of REST splicing attenuated SRRM4-induced STAT1 upregulation, suggesting that SRRM4 regulates STAT1 expression through REST splicing (Figure 5H & S19B). Consistently, STAT1 expression was significantly upregulated in REST splicing mutant mice (TAC-MT) compared with wild-type controls (TGC-WT), confirming that SRRM4-mediated REST splicing governs STAT1 expression (Figure 5I). As a transcriptional repressor, REST binds the RE-1 element of target genes to inhibit their transcription. The putative REST binding site in the STAT1 promoter was predicted by the JASPAR database and confirmed by ChIP and EMSA assays (Figure 5J-K). Dual luciferase reporter assays further showed that luciferase activity driven by the wild-type RE-1 sequence was significantly decreased by REST compared with mutant sequence groups (Figure 5L). ChIP assays showed that REST knockdown increased RNA polymerase binding at the STAT1 transcription initiation site, and RNA run-on assays indicated that REST upregulation inhibits STAT1 transcription initiation (Figure S19C-D). These data identify STAT1 as the direct trans-repression target of REST.

**Figure 5.**
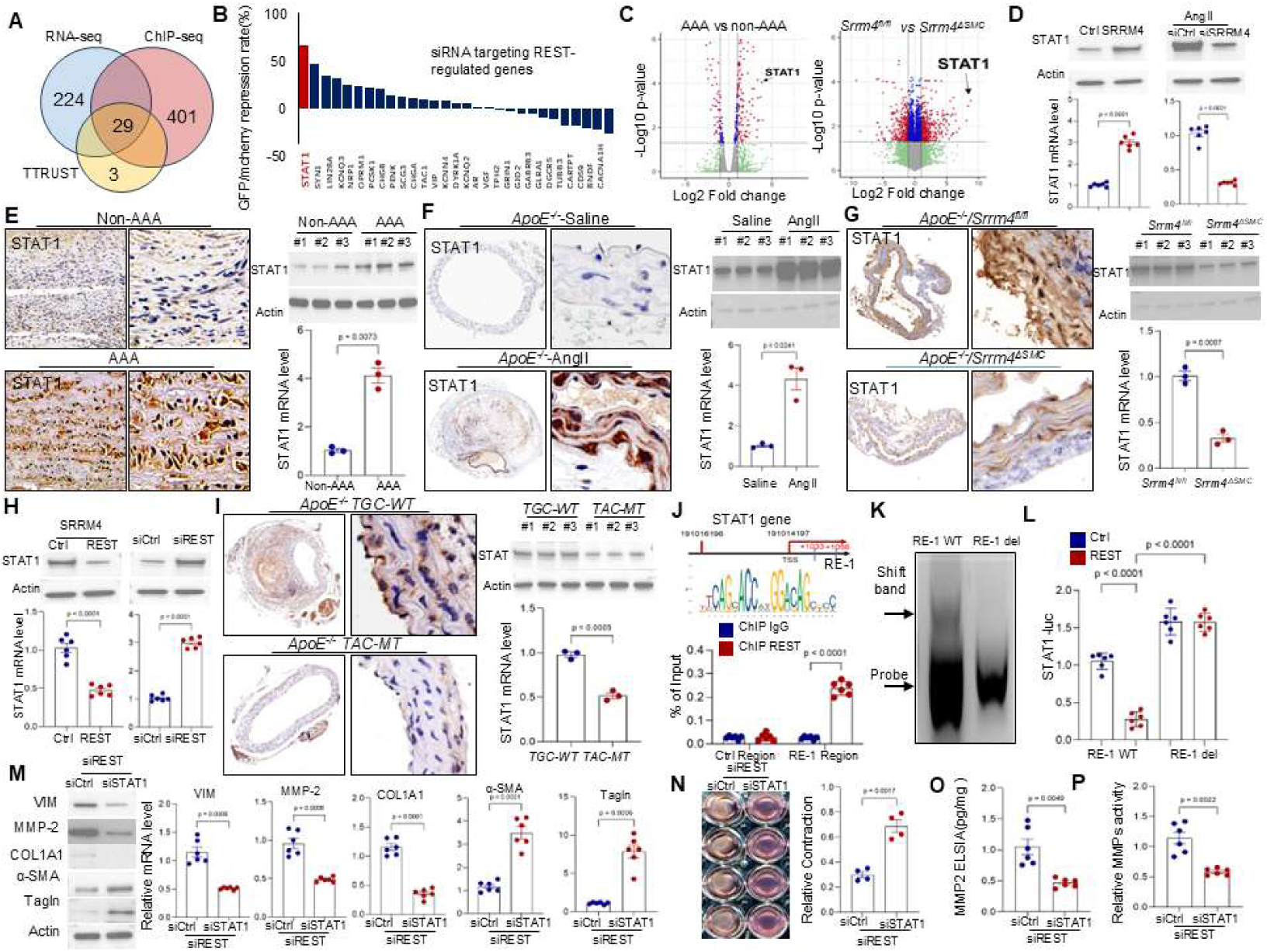
STAT1 is the direct trans-repression target of REST in regulating VSMC phenotype switch. **A)** VSMCs overexpressing ctrl or flag-tagged REST were used to extract total RNA for RNA-seq analysis of genes with significantly altered expression after REST overexpression. ChIP-seq was performed using flag antibodies to analyze REST-binding genes, and combined with the TTUST transcription factor database, a Venn diagram was used to show the 29 overlapped genes of the three groups. **B)** SRRM4 and the VSMC phenotype switch reporting system were transfected into VSMCs, followed by the transfection of siRNA targeting splicing sites of the 29 genes to inhibit their expression. The suppression rate of GFP protein fluorescence intensity was then measured. **C)** The volcano plot displays the differentially expressed genes in aortic tissues of AAA and non-AAA patients, as well as the differentially expressed genes in *ApoE^−/−^/Srrm4^flox/flox^* and *ApoE^−/−^/Srrm4^ΔSMC^* mice. **D)** Total RNA and protein of VSMCs overexpressing SRRM4, or VSMCs with silenced SRRM4 after Ang treatment were collected, STAT1 expression was detected using immunoblotting and real-time qPCR. **E-G)** Quantification of STAT1 expression were measured by realtime-qPCR and immunoblotting in E) human AAA and non-AAA segments, F) abdominal aortic segments derived from *ApoE^−/−^* mice treated with saline or AngII, G) abdominal aortas of *ApoE^−/−^/Srrm4^flox/flox^* and *ApoE^−/−^/Srrm4^ΔSMC^* mice, Representative images of STAT1 expression by IHC staining were shown. **H)** Total RNA and protein of VSMCs overexpressing REST, or VSMCs with silenced REST were collected, STAT1 expression was detected using immunoblotting and real-time qPCR. **I)** Quantification of STAT1 expression were measured by realtime-qPCR and immunoblotting in TGC-WT and TAC-MT mice treated with AngII. Representative images of STAT1 expression by IHC staining were shown. **J)** VSMCs were used to perform ChIP assays with the REST or IgG antibody. Eluted DNA fragments were used as templates for real-time qPCR to measure the enrichment of REST to RE-1 elements in the STAT1 promoter. Signals were calculated as percentage of input. **K)** Construct DNA probes with either the complete or deleted RE-1 element of the STAT1 gene and transfect them into VSMCs. The EMSA assay was performed as described in the material and method section. **L)** Luciferase reporter vectors with complete or deleted RE-1 element of the STAT1 gene were constructed and transfect into VSMCs. Relative luciferase activity was detected in VSMC after transfection with REST or control expression vectors. **M-P)** VSMCs with REST knocked down were treated with control siRNA or siRNA targeting STAT1 for 48 hours. M) Quantification of mRNA and protein levels of α-SMA, Tagnl, COL1A1, MMP2 and VIM in VSMCs; N) VSMCs contraction ability were measured by collagen gel contraction assay; O) Secreted MMP2 levels were measured by ELISA; P) MMP activity of indicated cells were measured by MMP activity assays.

STAT1 functions as a transcription factor. Knocking down STAT1 ameliorated VSMC phenotype switching and MMP2 expression induced by REST loss, whereas STAT1 overexpression promoted VIM, MMP2, and COL1A1 expression (Figure 5M-P and S20A), suggesting that STAT1 promotes VSMC phenotype switching. Since the Myocardin/SRF complex maintains the VSMC contractile phenotype, we performed Co-IP assays and found that STAT1 binds Myocardin and disrupts Myocardin/SRF complex formation (Figure S20B-C). ChIP assays further showed that STAT1 binds the promoter regions of MMP2 and VIM, while simultaneously inhibiting SRF binding to the α-SMA and Tagln promoters (Figure S20D-E). Thus, STAT1 orchestrates VSMC phenotype switching through both direct activation of synthetic genes and indirect repression of contractile genes (Figure S20F). Mechanistically, these data identify STAT1 as the direct trans-repression target of REST, mediating SRRM4/REST axis-induced VSMC phenotype switching.

### Targeting STAT1 protects against AAA formation in animal model

To further investigate the role of STAT1 in AAA pathogenesis *in vivo*, we constructed the VSMC-specific *Stat1* knockout *ApoE^−/−^* mice (*ApoE^−/−^*/*Stat1^ΔSMC^*) to generate AngII-induced AAA animal model (Figure 6A). Silencing of STAT1 in VSMC resulted in a marked reduction in AngII-induced AAA progression, as demonstrated by significant differences in AAA incidence, aortic diameter and elastin degradation between *ApoE^−/−^*/*Stat1^ΔSMC^* and *ApoE^−/−^*/*Stat1^flox/flox^* groups (Figure 6B-G). While STAT1-inhibiting therapies have been extensively used for hematological malignancies, their potential to inhibit AAA progression has not been investigated. We therefore examined whether STAT1 inhibition could attenuate VSMC phenotype switching and AAA progression. Fludarabine has been reported to specifically suppress STAT1 without affecting other STAT family members. ApoE-/-mice were subjected to AAA induction followed by fludarabine treatment, and phosphorylated STAT1 levels in aortic tissue were measured to confirm the inhibitory effect of fludarabine on STAT1 activity (Figure 6H). As expected, fludarabine-treated mice showed less AAA incidence and repressed aortic dilation than vehicle-treated mice (Figure 6I-N), with no significant differences in systolic/diastolic blood pressure, heart rate, or plasma lipids (Figure S21). To assess the dependency of fludarabine’s cellular effects on STAT1, AngII-stimulated VSMCs were treated with siSTAT1, fludarabine, or both. Fludarabine alone suppressed AngII-induced upregulation of COL1A1, MMP2, and VIM, whereas it produced no additional suppression in siSTAT1-treated VSMCs (Figure S22A). Conversely, STAT1 overexpression significantly restored the expression of these synthetic markers suppressed by fludarabine (Figure S22B), demonstrating that the inhibitory effect of fludarabine on VSMC phenotype switching is largely STAT1-dependent. To evaluate whether this protection involves immune modulation, we examined peripheral blood counts and aortic immune cell infiltration. Aortic tissue sections showed no significant differences in F4/80+ macrophage or MPO+ neutrophil infiltration between fludarabine- and vehicle-treated groups (Figure S22C), indicating that fludarabine protects against AAA primarily through direct STAT1 inhibition in VSMCs rather than through global immunosuppression. Collectively, these findings suggest that targeting STAT1 provides protection against AAA progression, offering a potential therapeutic avenue for this vascular pathology.

**Figure 6.**
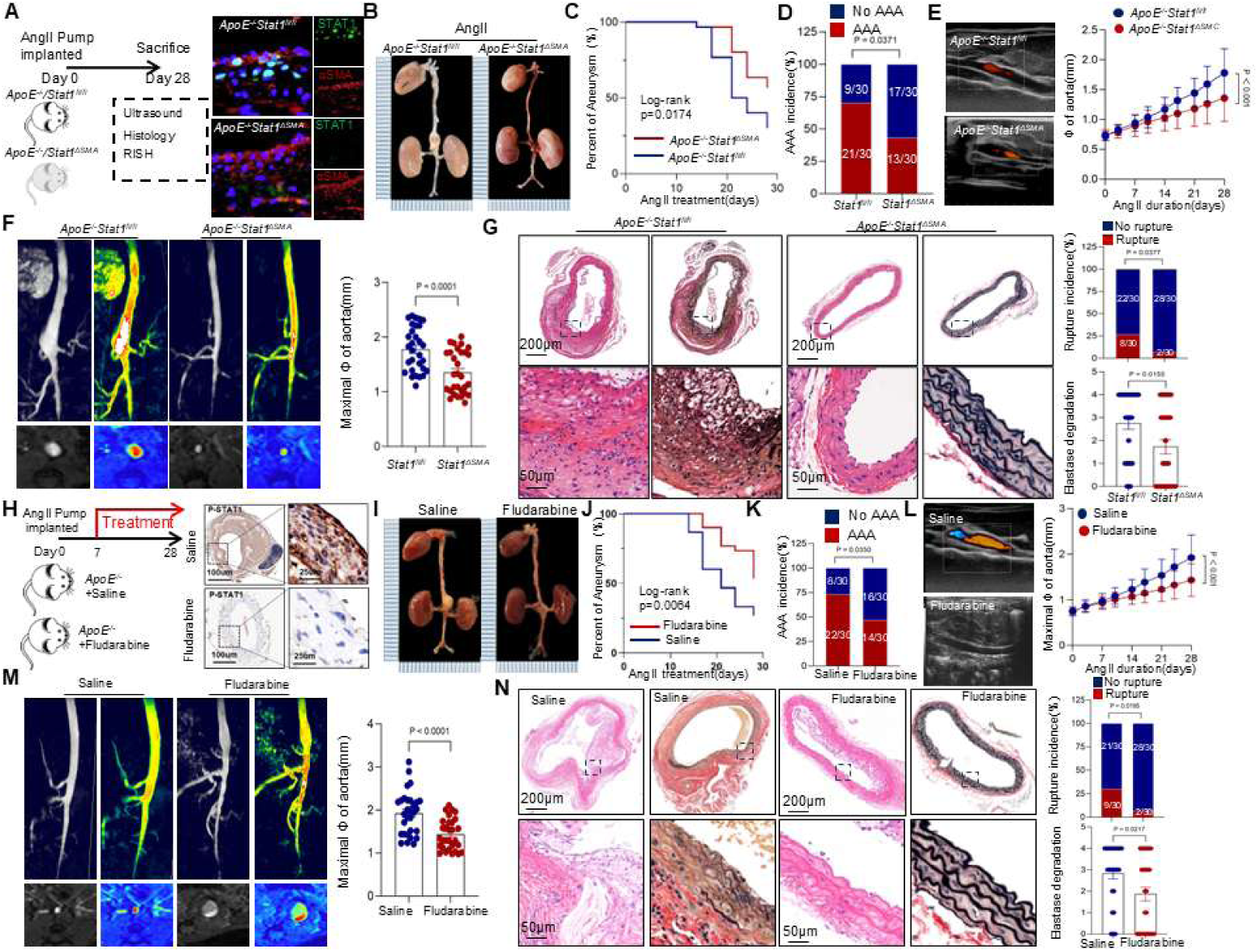
STAT1 inhibition protects against AngII-induced AAA formation. **A)** Schematic protocol of: *ApoE^−/−^*/*Stat1^flox/flox^* and *ApoE^−/−^*/*Stat1^ΔSMC^* mice were subcutaneously injected with saline or AngII by a mini osmotic pump for 28 days. **B)** Representative images of the macroscopic features of AAA formation in indicated groups. **C)** Survival curves in indicated groups. Survival data were analyzed by the Kaplan-Meier method and compared using log-rank tests. **D)** The incidence of AngII-induced aneurysm in indicated groups was analyzed by the Fisher exact test. **E-F)** Quantification of the maximal diameter of suprarenal abdominal aortas in indicated groups. Representative images of the abdominal aorta visualized by MRI and B-mode ultrasound with duplex in the indicated groups. Growth curves of suprarenal abdominal aortic diameter measured by MRI and B-mode ultrasound in the indicated groups. **G)** Representative H&E and EVG staining images of suprarenal abdominal aortas in indicated groups after 21 days of saline or fludarabine infusion. The incidence of AngII-induced aneurysm in indicated groups was analyzed by the Fisher exact test. Quantification of elastin degradation by counting the number of breaks per vessel in suprarenal aortic segments, based on EVG staining. **H)** Schematic protocol of: *ApoE^−/−^* mice were subcutaneously injected with AngII by a mini osmotic pump and treated with either Saline or Fludarabine 50ug/g body weight for 21 days. IHC was used to verify the inhibition of STAT1 in smooth muscle cells. **I)** Representative images of the macroscopic features of AAA formation in indicated groups. **J)** Survival curves in indicated groups. Survival data were analyzed by the Kaplan-Meier method and compared using log-rank tests. **K)** The incidence of AngII-induced aneurysm in indicated groups was analyzed by the Fisher exact test. **L-M)** Quantification of the maximal diameter of suprarenal abdominal aortas in indicated groups. Representative images of the abdominal aorta visualized by MRI, and by B-mode ultrasound with duplex analysis, in the indicated groups. Growth curves of suprarenal abdominal aortic diameter measured by MRI and B-mode ultrasound in the indicated groups. N) Representative H&E and EVG staining images of suprarenal abdominal aortas in indicated groups after 21 days of saline or fludarabine infusion. The incidence of AngII-induced aneurysm in indicated groups were analyzed by the Fisher exact test. Quantification of elastin degradation by counting the number of breaks per vessel in suprarenal aortic segments, based on EVG staining.

## DISCUSSION

As population aging progresses, AAA is expected to become more prevalent and result in an increasing number of medical emergencies. A better understanding of the molecular mechanisms underlying AAA development is therefore necessary for designing therapeutic strategies. In this study, we elucidated a previously unrecognized role of the splicing factor SRRM4 in AAA etiology, providing new insights into its pathogenesis. Our data reveal that SRRM4 drives alternative splicing of the REST gene to promote VSMC phenotypic switching and MMP expression, ultimately promoting AAA formation, development, and rupture. Importantly, targeting the SRRM4/REST/STAT1 axis effectively counteracted the adverse effects of SRRM4 on VSMC phenotypic switching and attenuated AAA formation. This study illustrates the critical role of SRRM4 in AAA and highlights the mechanistic potential for AAA prevention.

Serine/arginine-rich proteins are transcription factors that play a key role in regulating gene expression in both physiological and pathological contexts, including cancer and cardiovascular disease^14, 27–30^. SRRM4, a distinctive member of the SR protein family, is recognized as a neural-specific key regulator in the development of the neural system^31^. Beyond its traditional role in neural cell differentiation, recent studies have implicated SRRM4 in the pathogenesis of cancer and hearing loss^32–34^. However, its function in cardiovascular physiology and pathology has remained undefined. Our findings uniquely identify SRRM4 upregulation in VSMCs of AAA tissues (Figure 1 and S1-4), and demonstrate protection against AAA in both AngII-induced and CaCl2-induced models with VSMC-specific SRRM4 knockout (Figure 2 and S8). The detrimental role of SRRM4 was further confirmed by *in vitro* experiments showing SRRM4-induced VSMC phenotype switching and MMP2 activation (Figure 3). The specificity of SRRM4 upregulation in VSMCs, but not in endothelial or myeloid cells, highlights its unique role in mediating VSMC phenotypic switching, a critical event in AAA pathogenesis (Figure S9). Crucially, the protective phenotype was consistently reproduced across three independent VSMC-targeting systems: constitutive Tagln-Cre-mediated knockout, DOX-inducible knockdown, and tamoxifen-inducible Tagln-CreERT2-mediated knockout (Figure 2 and Figure S10). This convergence of evidence from orthogonal genetic approaches firmly establishes the VSMC-intrinsic role of SRRM4 in AAA pathogenesis.

Our data reveal that SRRM4 compromises the function of REST, a transcriptional repressor essential in vascular pathology. Loss of REST has been reported to drive VSMC acquisition of synthetic and proliferative characteristics. Previous research has shown that REST is post-translationally modulated by β-TRCP during VSMC phenotype transition.^21, 35^. Our findings extend this knowledge by demonstrating that SRRM4 directly inhibits REST through alternative splicing, generating the non-functional REST4 isoform, which in turn promotes VSMC phenotype switching and AAA development (Figure 3–4 and S11-18). Targeting STAT1, a downstream component of the SRRM4/REST axis, effectively attenuated AAA formation and counteracted the adverse effects of SRRM4 overexpression (Figure 6 and S19-20).

While the REST/STAT1 axis represents the dominant effector pathway, other SRRM4 targets also contribute to the full spectrum of pathology. For example, SRRM4 promotes alternative splicing of Abi1(Figure S3), resulting in upregulation of macrophage-like markers in SMCs, consistent with our previous findings^25^. Thus, we propose a hierarchical model in which REST/STAT1 serves as the primary node controlling VSMC contractile-to-synthetic switching, while additional targets fine-tune secondary pathological features. Intervening at this downstream node, whether by blocking REST splicing, restoring REST function, or inhibiting STAT1, is sufficient to reverse the major phenotypic consequences of SRRM4 activation, even when other SRRM4-dependent splicing events remain unaltered.

Understanding the regulatory interplay between SRRM4, REST splicing, and STAT1 activation is essential for developing targeted therapies to halt or reverse AAA progression. Our results demonstrated that targeting STAT1 significantly mitigates AAA progression, as shown in VSMC-specific Stat1 knockout mice and pharmacological intervention with fludarabine, which has been reported to suppress STAT1 activation. These interventions not only reduce AAA incidence and progression but also reverse VSMC phenotypic switching (Figure 6 and S22). These findings suggest that targeting the SRRM4/REST/STAT1 pathway offers a novel therapeutic strategy for AAA. This study enriches the understanding of AAA pathogenesis by introducing novel molecular targets, moving beyond current surgical and endovascular limitations toward more nuanced molecular management.

In conclusion, our investigation sheds light on the critical role of SRRM4 in AAA pathogenesis and highlights the therapeutic potential of targeting the SRRM4/REST/STAT1 pathway. Given the lack of effective pharmacological treatments for AAA, the elucidation of this novel pathway may contribute to innovative treatment strategies that could improve patient outcomes.

## Data Availability

All data generated or analyzed during this study are included in this published article and its supplementary information files.

## Acknowledgments

We are also particularly thankful to Professor Wei Zhang from Fudan University for his valuable support and expert guidance in statistical analysis.

---

**Figure S1.**
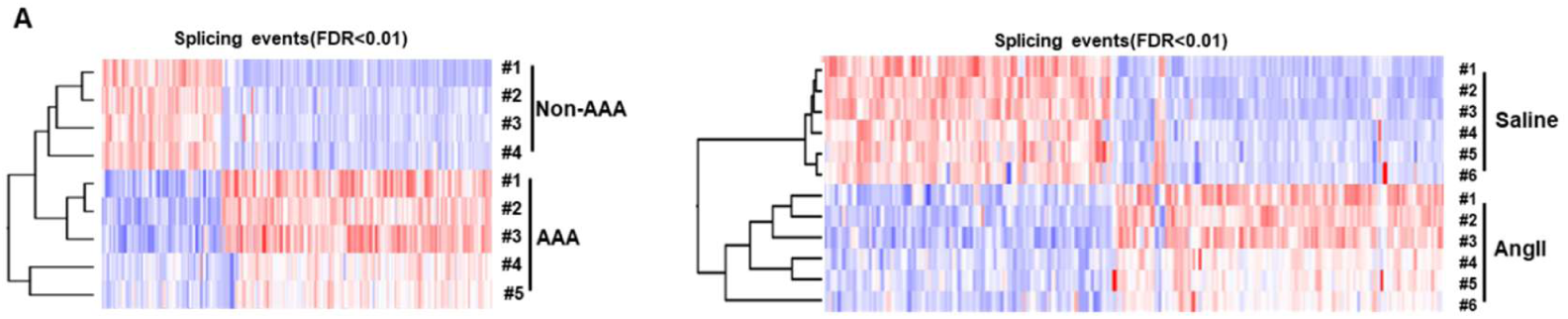
**A)** AAA-specific alternative splicing signatures are generated based on 106 differential AS events between AAA and non-AAA samples from the patient cohort and animal models

**Figure S2.**
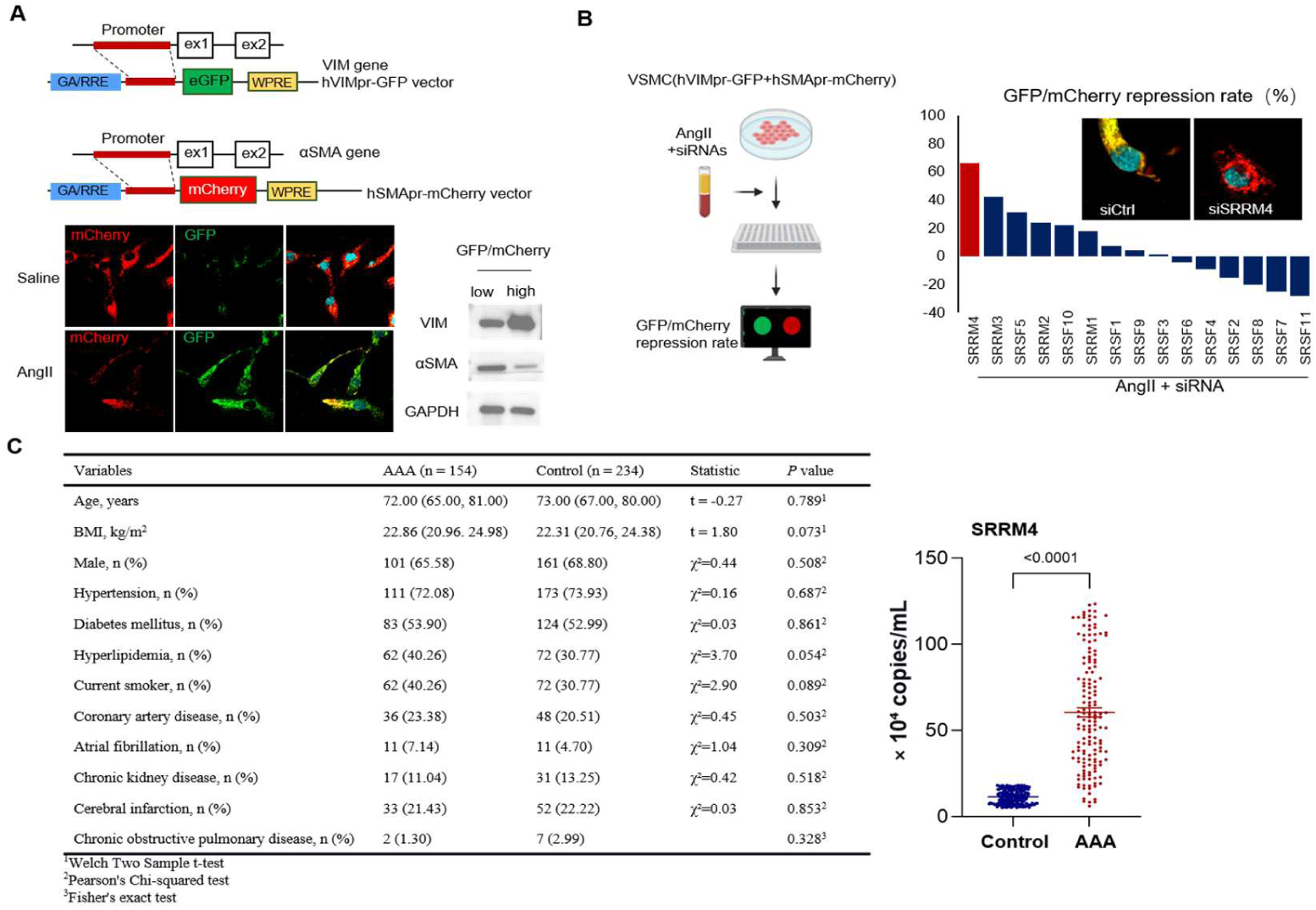
**A)** A schematic diagram shows the structure of VSMC phenotype switch reporting system. Efficacy of reporting system was validated by immunoblotting. A series of siRNAs targeting SR superfamily members were transfected into VSMC report system treated with AngII. eGFP/mCherry intensity were measured and repression rates were calculated. The siRNA targeting SRRM4 showed highest repression rate. B) Baseline characteristics of the study participants in the exosome cohort. C) Plasma exosomes were isolated from 154 AAA patients and 234 age- and sex-matched non-AAA controls. Relative SRRM4 mRNA levels in exosomes were quantified by RT-qPCR

**Figure S3.**
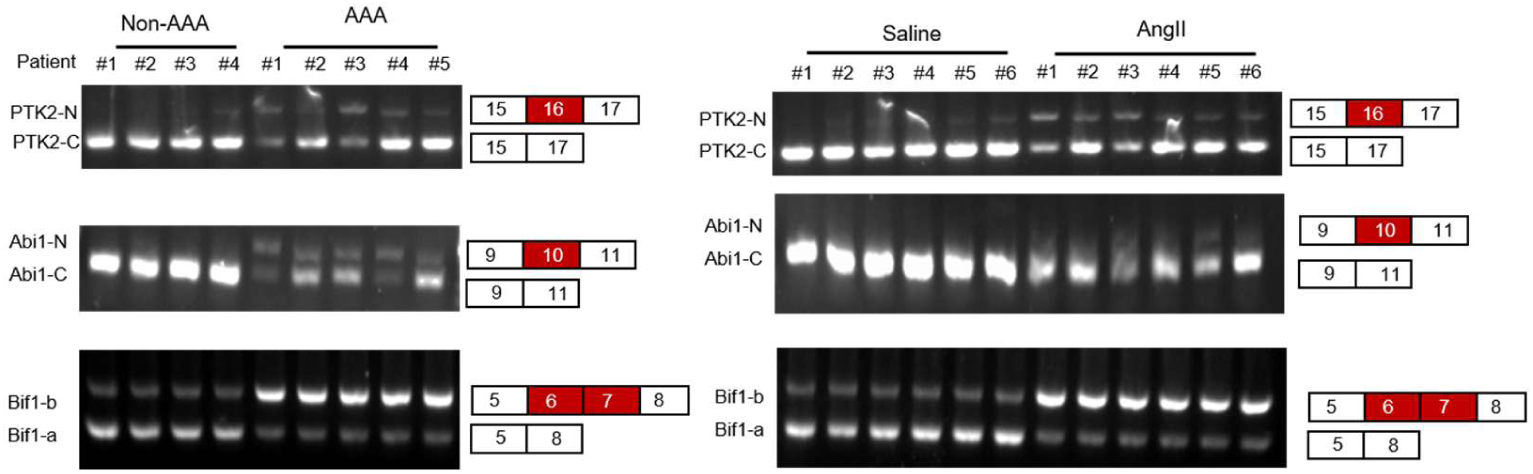
**A)** Total RNA of the primary cultured VSMC derived from human AAA and non-AAA segments and mouse abdominal aortic segments were collected. Alternative splicing events of SRRM4-target genes were validated by RT-PCR.

**Figure S4.**
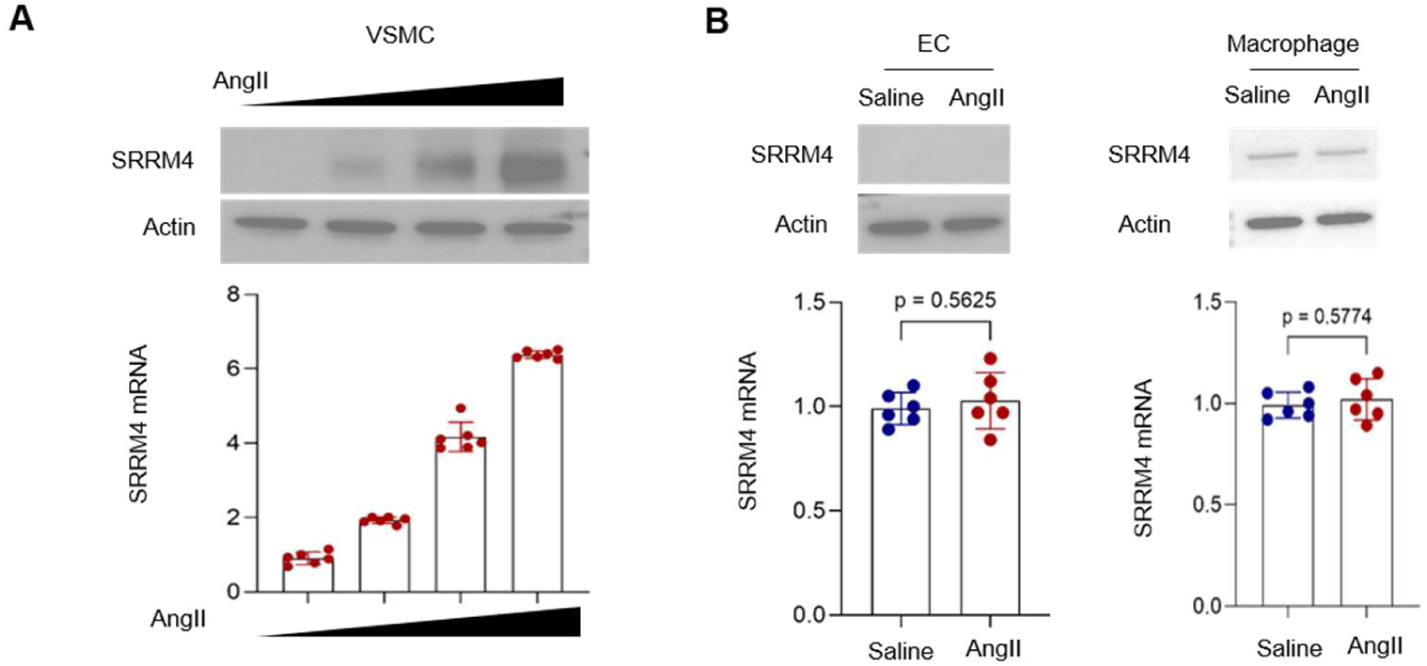
**A)** VSMC were treated with AngII for 7 days, expression of SRRM4 mRNA and protein were measured by realtime-qPCR and immunoblotting. **B**) Primary cultured endothelial and macrophage derived from mouse abdominal aortic segments were stimulation with AngII for 48 hours, expression of *Srrm4* mRNA and protein were measured by realtime-qPCR and immunoblotting.

**Figure S5.**
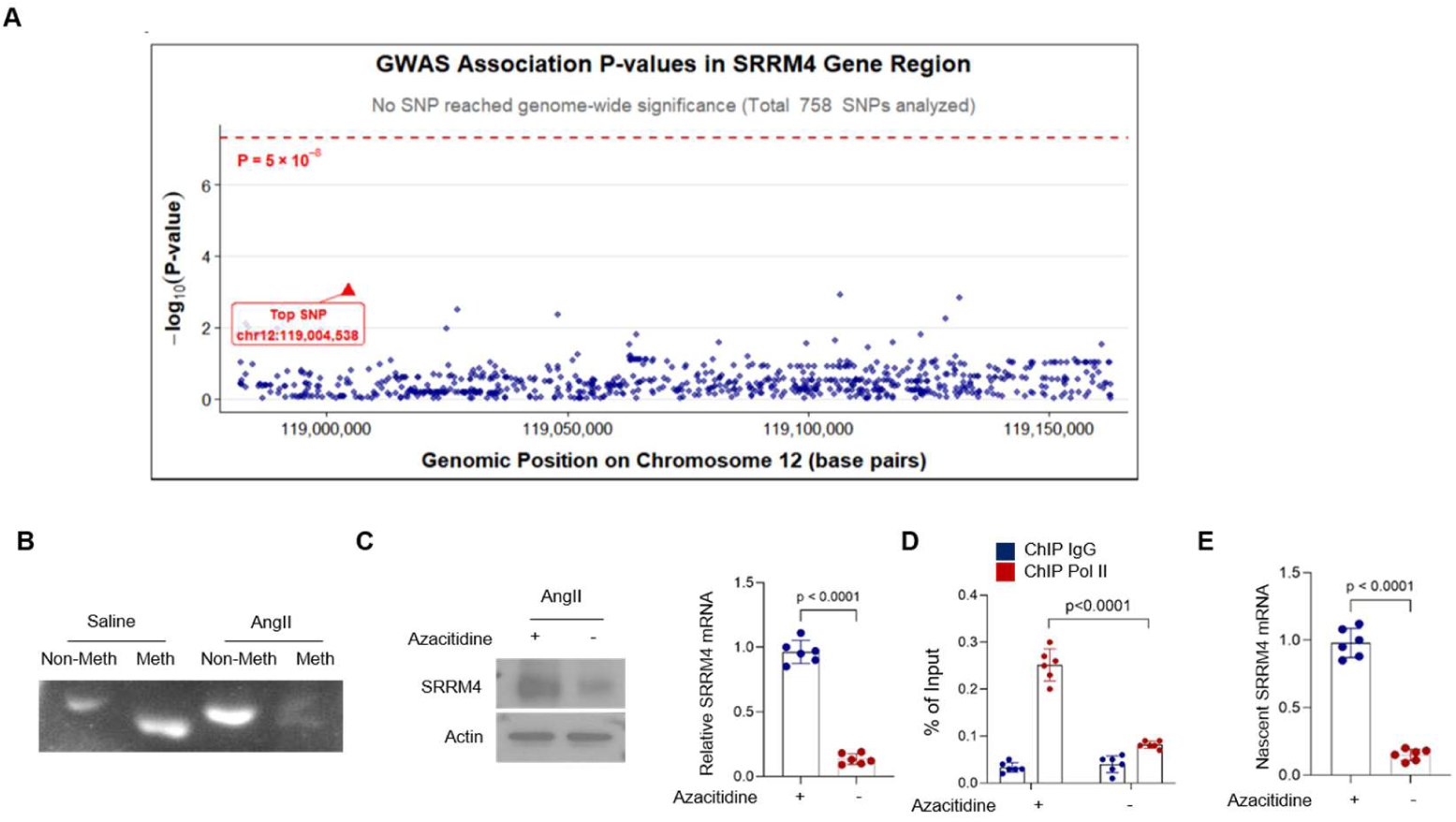
**A)** Each point represents a single nucleotide polymorphism (SNP) within the SRRM4 genomic region (chr12:118,981,541-119,163,051, GRCh37/hg19). A total of 758 SNPs were analyzed, none of which reached genome-wide significance. The top-associated SNP (chr12:119,004,538, p = 9.81 × 10⁻⁴) is highlighted in red. The absence of points above the significance threshold indicates a lack of significant association between common genetic variants in the SRRM4 region and AAA risk. GWAS data were obtained from the IEU OpenGWAS database (study ID: GCST90018563). **B)** Total DNA of the primary cultured VSMC derived from mouse abdominal aortic segments were collected for DNA methylation analyze as described in the M&M section. **C-E)** VSMC were treated with AngII plus saline or DNA methyltransferase inhibitor Azacitidine for 24 hours. B) Total protein and RNA were collected and expression of SRRM4 were detected by immunoblotting and realtime-qPCR, C) Total DNA were used to perform Ch-IP assays with the RNA polymerase II antibodies. Eluted DNA fragments were used as templates for real-time qPCR to measure the enrichment of RNA polymerase II to SRRM4 gene, Signals were calculated as the percentage of input, **D)** Cells were used to perform RNA run-on assays. Nascent SRRM4 mRNAs were measured by real-time qPCR.

**Figure S6.**
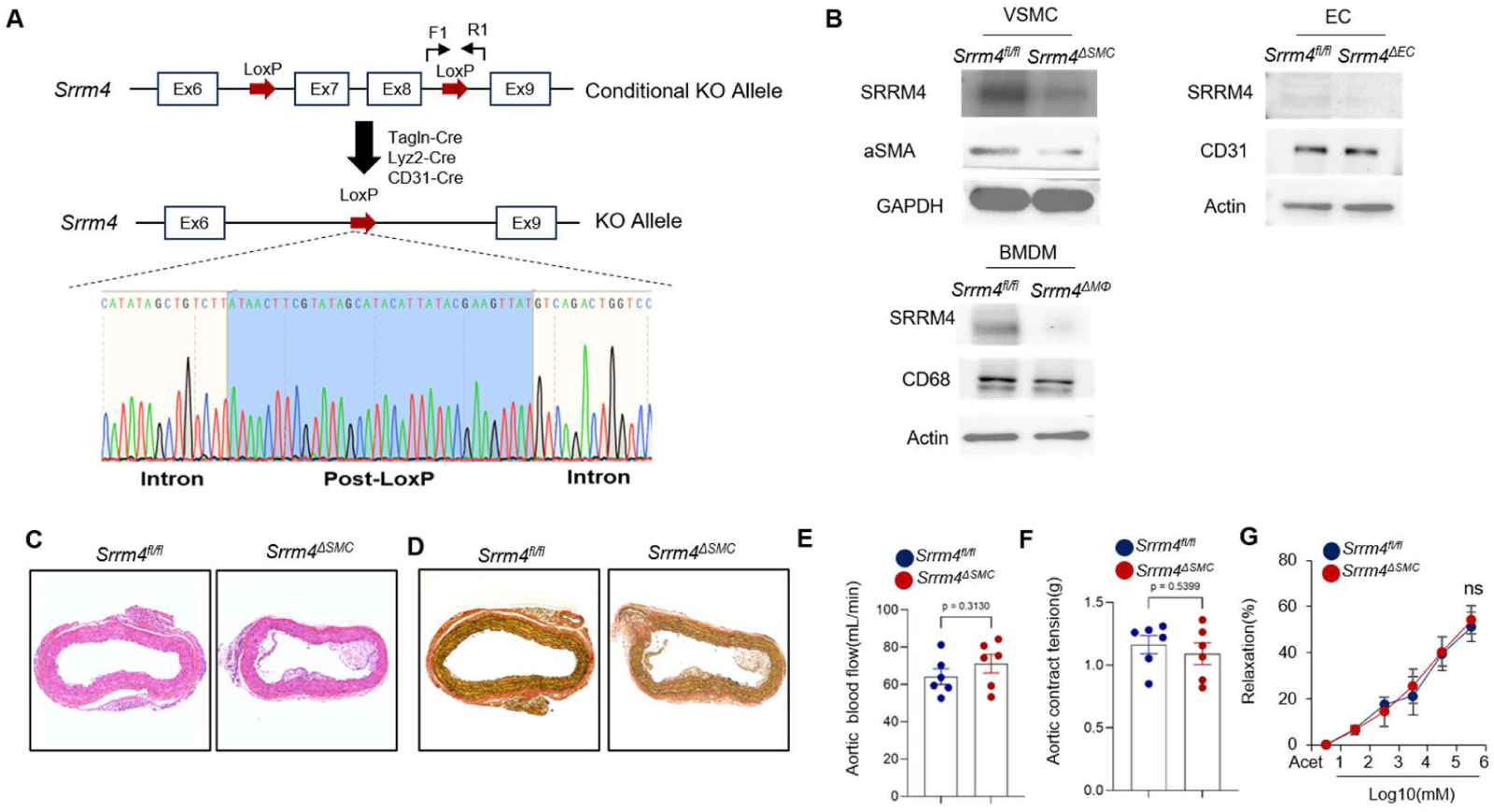
**A)** Schematic diagram of the construction of conditional VSMC specific Srrm4 knockout mice. Validation of CRISPR efficacy was confirmed by Sanger sequencing. **B)** Validation of the effective inactivation of Srrm4 in VSMC from different mouse strains by immunoblotting. **C-D)** Representative C) H&E and D) EVG staining images of suprarenal abdominal aortas in indicated groups. **E)** Aortic blood flow in each group were monitored by ultrasound. Quantitative data are presented. **F-G)** Functional analysis of harvested IVC segments assessing F) contraction responses to 40mM KCl and G) Relaxation responses to the cumulative addition of acetylcholine.

**Figure S7.**
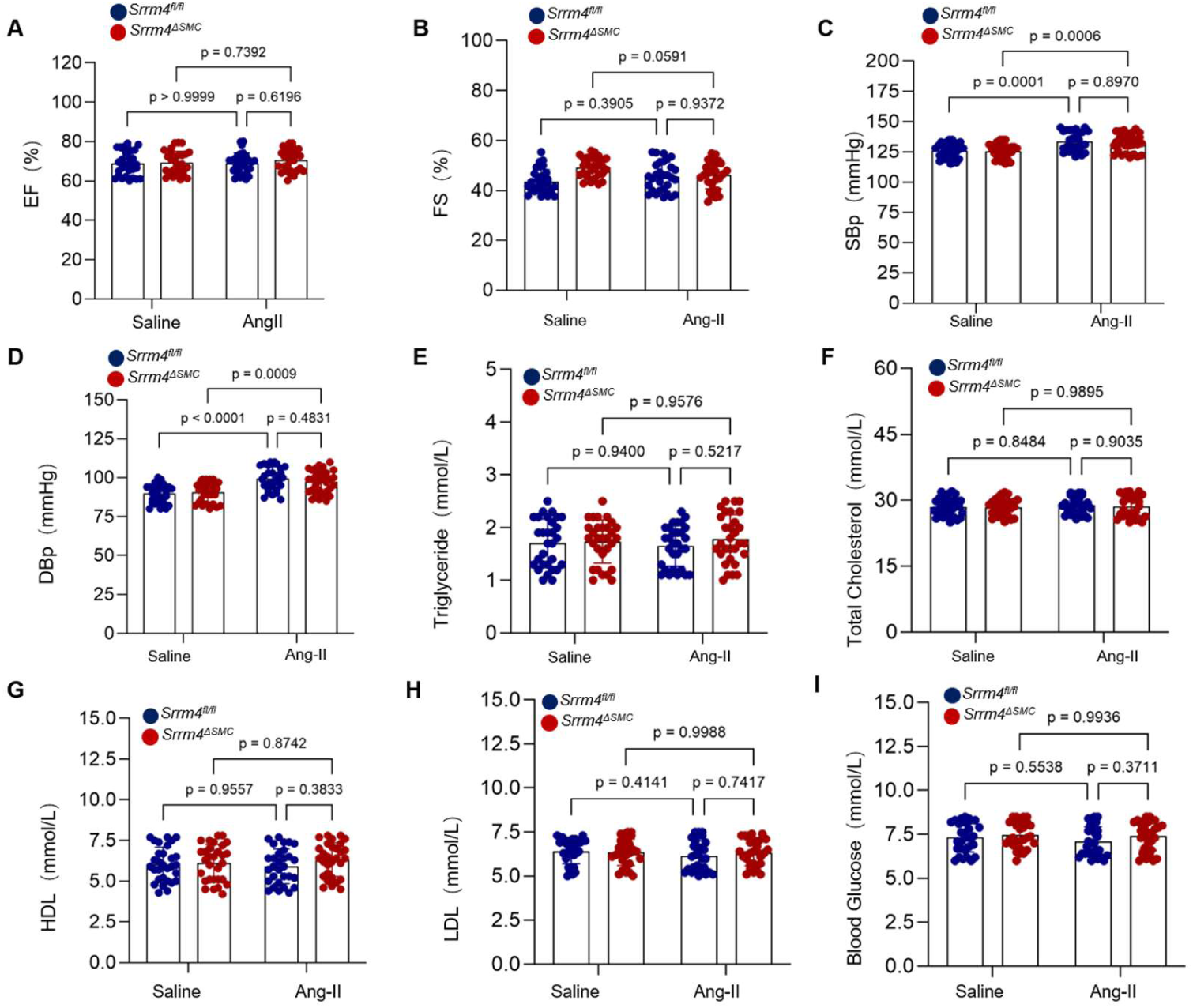
VSMC specific *Srrm4* knockout did not overtly affect cardiac function, blood pressure, and metabolic parameters of *ApoE*^−/−^.

**Figure S8.**
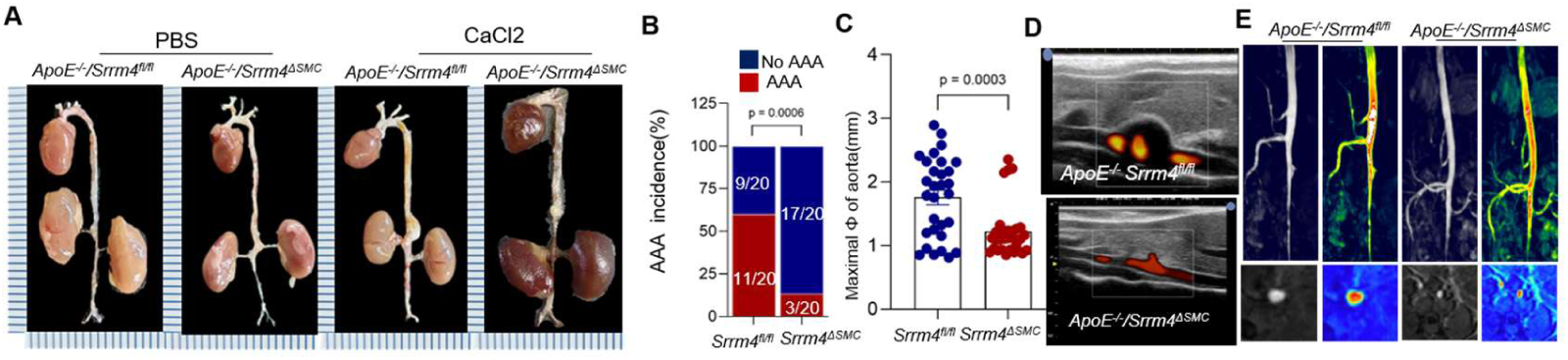
**A))** Representative images showing the macroscopic features of aortas from PBS-or CaCl_2_-treated *ApoE*^−/−^/*Srrm4^flox/flox^*and *ApoE*^−/−^/*Srrm4^ΔSMC^* mice. **B)** The incidence of aneurysm in indicated groups were analyzed by the Fisher exact test. **C)** Quantification of the maximal diameter of suprarenal abdominal aortas from indicated groups. **D-E)** Representative images of the abdominal aorta visualized by B-mode ultrasound with duplex analysis and by MRI in the indicated groups.

**Figure S9.**
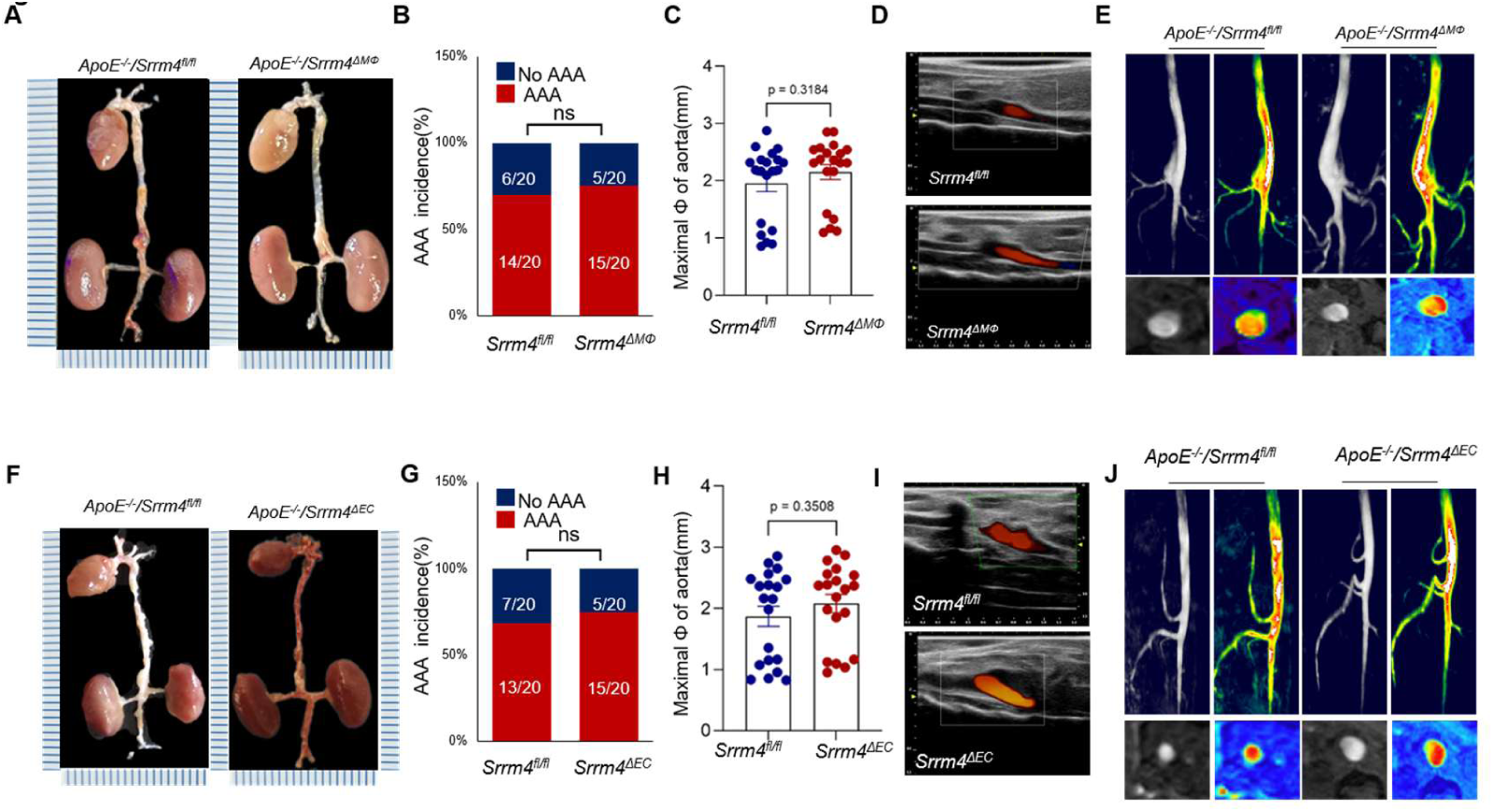
**A)** Representative images showing the macroscopic features of aortas from Saline-or AngII-treated *ApoE*^−/−^/*Srrm4^flox/flox^* and *ApoE*^−/−^/*Srrm4^ΔMΦ^* mice mice. **B)** The incidence of aneurysm in indicated groups were analyzed by the Fisher exact test. **C)** Quantification of the maximal diameter of suprarenal abdominal aortas from indicated groups. **D-E)** Representative images of the abdominal aorta visualized by B-mode ultrasound with duplex analysis and by MRI in the indicated groups. **F)** Representative images showing the macroscopic features of aortas from Saline-or AngII-treated *ApoE*^−/−^/*Srrm4^flox/flox^* and *ApoE*^−/−^/*Srrm4^ΔEC^* mice mice. **G)** The incidence of aneurysm in indicated groups were analyzed by the Fisher exact test. **H)** Quantification of the maximal diameter of suprarenal abdominal aortas from indicated groups. **I-J)** Representative images of the abdominal aorta visualized by B-mode ultrasound with duplex analysis and by MRI in the indicated groups.

**Figure S10.**
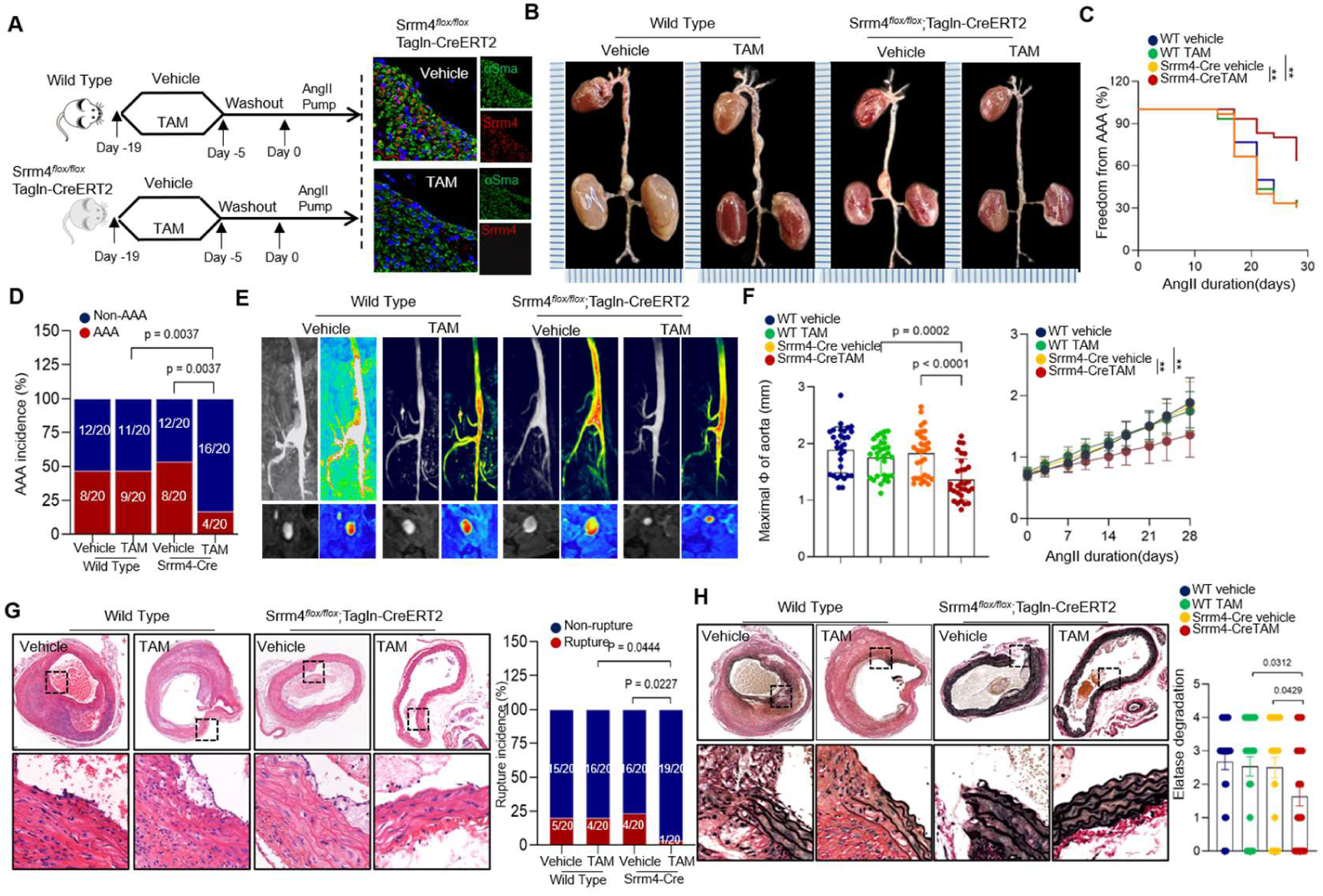
**A)** The transgenic mice with smooth muscle cell-specific knockout of the SRRM4 protein induced by tamoxifen(TAM) were constructed as described in the Material and Methods. Immunofluorescence was used to verify the specific expression of the SRRM4 protein in smooth muscle cells. **B)** Representative images of the macroscopic features of AAA formation in indicated groups. **C)** Survival curves in indicated groups. Survival data were analyzed by the Kaplan-Meier method and compared using log-rank tests. **D)** The incidence of AngII-induced aneurysm in indicated group was analyzed by the Fisher exact test. **E)** Representative MRI images of the abdominal aorta in the indicated groups. Growth curves showing the diameter of the suprarenal abdominal aorta measured by MRI in the indicated groups. **F)** Quantification of the maximal diameter of suprarenal abdominal aortas from indicated groups. **G-H)** Representative H&E and EVG staining images of suprarenal abdominal aortas in indicated groups after 28 days of saline or AngII infusion. Quantification of elastin degradation by counting the number of breaks per vessel in suprarenal aortic segments, based on EVG staining. The rupture incidence of AngII-induced aneurysm in indicated group was analyzed by the Fisher exact test.

**Figure S11.**
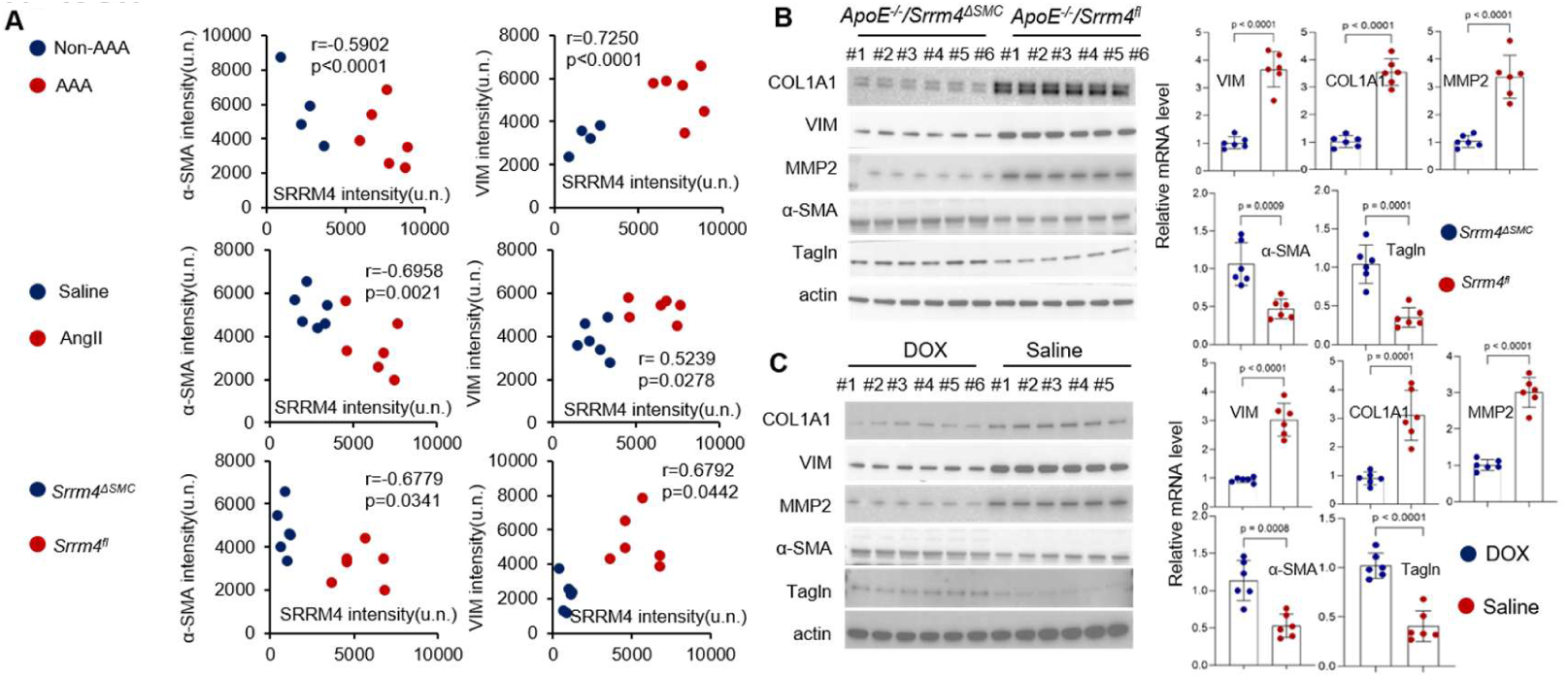
**A)** IF staining using SRRM4, α-SMA, and VIM antibodies were performed on human AAA and non-AAA segments, *ApoE*^−/−^/*Srrm4^flox/flox^* and *ApoE^−/−^*/*Srrm4^ΔSMC^* mice, as well as suprarenal abdominal aortas derived from TRE3G-Mouse SRRM4-CaG-Tet3G treated with saline or DOX, Pearson correlation of α-SMA and VIM with SRRM4 were calculated and plotted. **B-C)** Quantification of α-SMA, Tagln, COL1A1, VIM, and MMP2 mRNA and protein expression measured by realtime-qPCR and immunoblotting in mouse abdominal aortic segments derived from *ApoE*^−/−^/*Srrm4^flox/flox^* and *ApoE^−/−^*/*Srrm4^ΔSMC^* mice, as well as Tagln-rtTA; TRE3G-shRNA(Srrm4) mice treated with saline or DOX.

**Figure S12.**
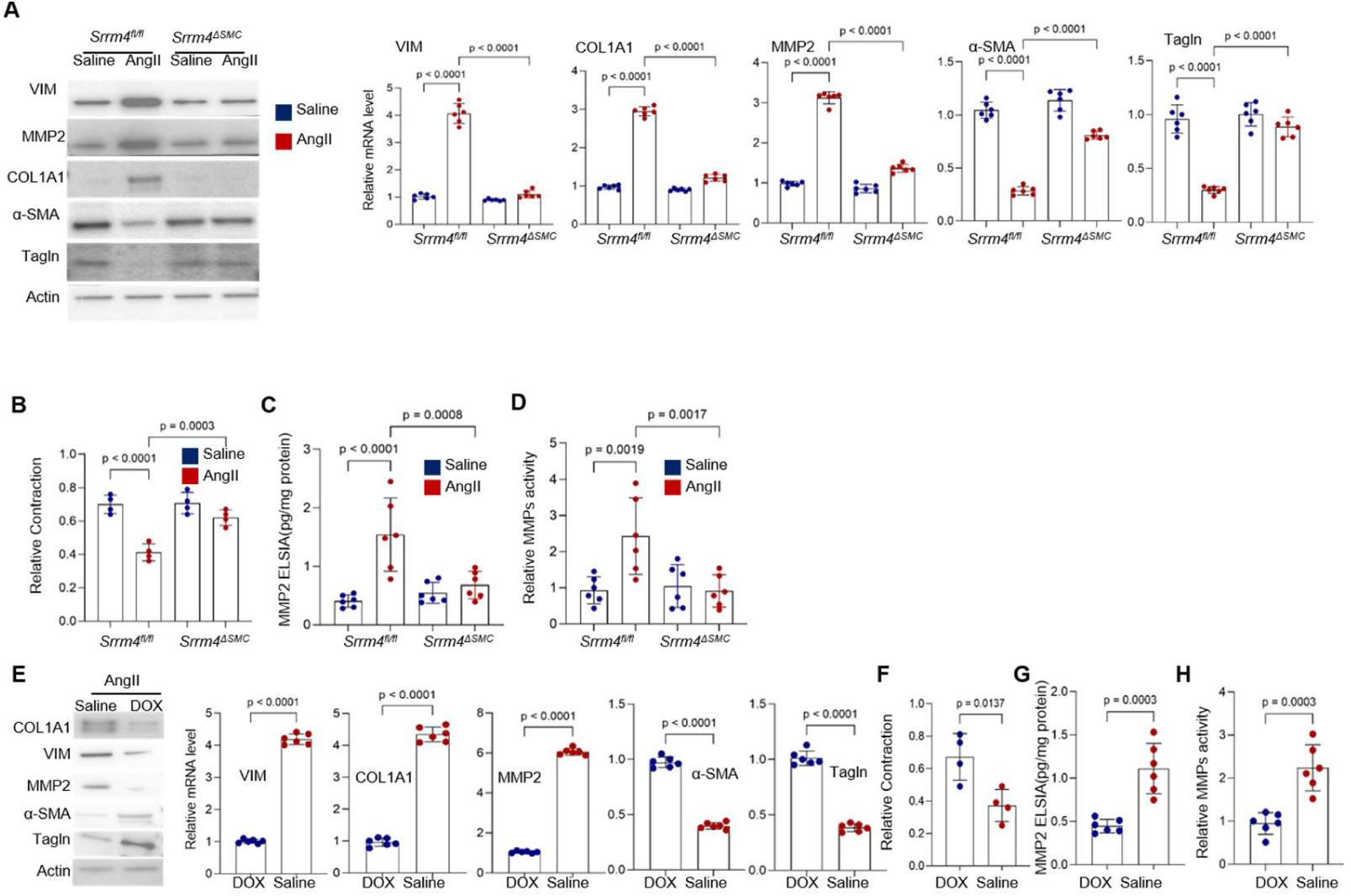
**A-D)** Primary cultured VSMC derived from Saline-or AngII-treated *ApoE*^−/−^/*Srrm4^flox/flox^* and *ApoE*^−/−^/*Srrm4^ΔSMC^* mice were collected, A)Total RNA and protein of α-SMA, Tagln, COL1A1, VIM, and MMP2 were detected by realtime-qPCR and immunoblotting, B) SMC contraction ability were measured by collagen gel contraction assay, C) Secreted MMP2 level were measured by ELISA, D) MMP activity of indicated cells were measured by MMP activity assays. **E-H**) The Primay culture VSMC derived from *Tagln-rtTA; TRE3G-shRNA(Srrm4)* mice were treated with Saline of DOX for 7 days, E) The total RNA and protein were collected and expression of α-SMA, Tagln, COL1A1, VIM, and MMP2 were detected by realtime-qPCR and immunoblotting. F) VSMCs contraction ability were measured by collagen gel contraction assay; G) Secreted MMP2 levels were measured by ELISA; H) MMP activity of indicated cells were measured by MMP activity assays.

**Figure S13.**
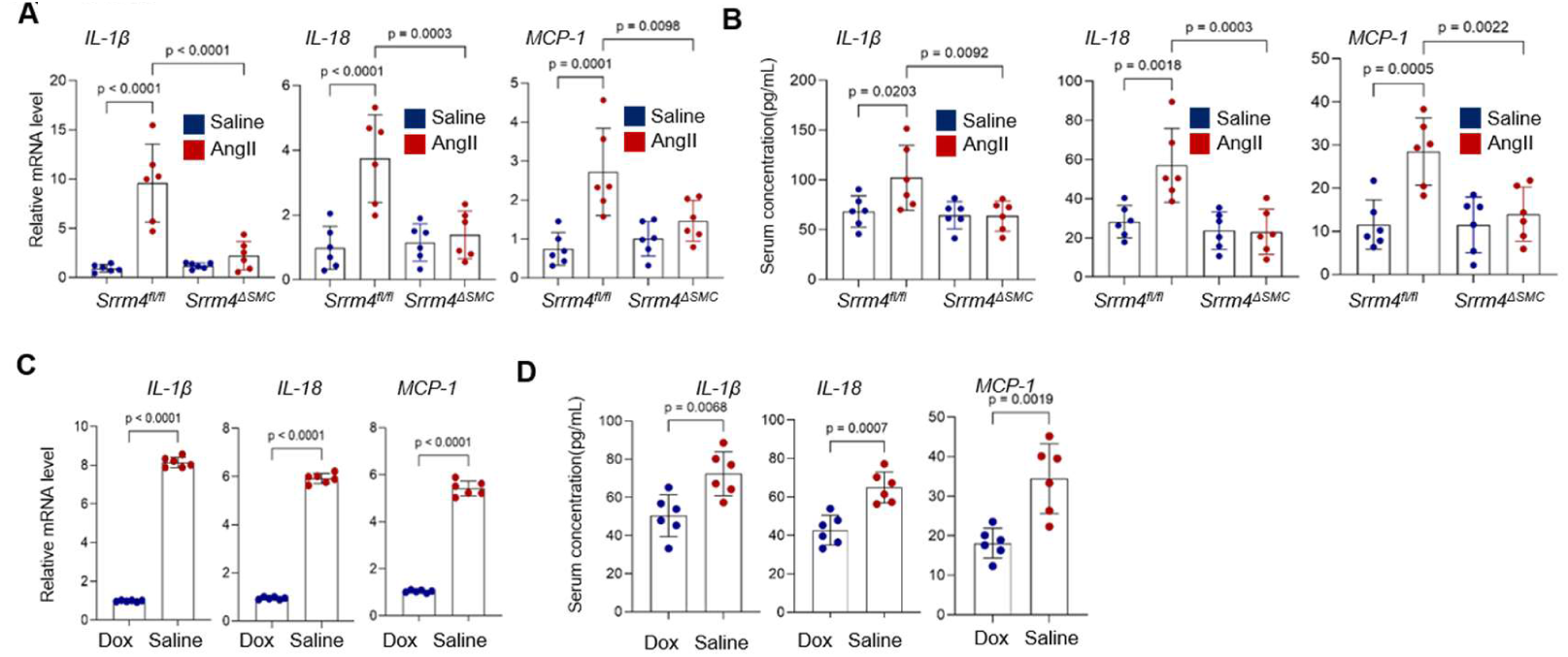
**A-B)** Primary cultured VSMC derived from Saline-or AngII-treated *ApoE*^−/−^/*Srrm4^flox/flox^* and *ApoE*^−/−^/*Srrm4^ΔSMC^* mice were collected. A) Intercellular levels of Il-1β, Il-18, and MCP-1 were detected by realtime-qPCR and immunoblotting, **B)** Secreted levels of Il-1β, Il-18, and MCP-1 were detected ELSIA. **C-D**) The Primay culture VSMC derived from *Tagln-rtTA; TRE3G-shRNA(Srrm4)* mice were treated with Saline of DOX for 7 days. C) Intercellular levels of Il-1β, Il-18, and MCP-1 were detected by realtime-qPCR and immunoblotting, D) Secreted levels of Il-1β, Il-18, and MCP-1 were detected ELSIA.

**Figure S14.**
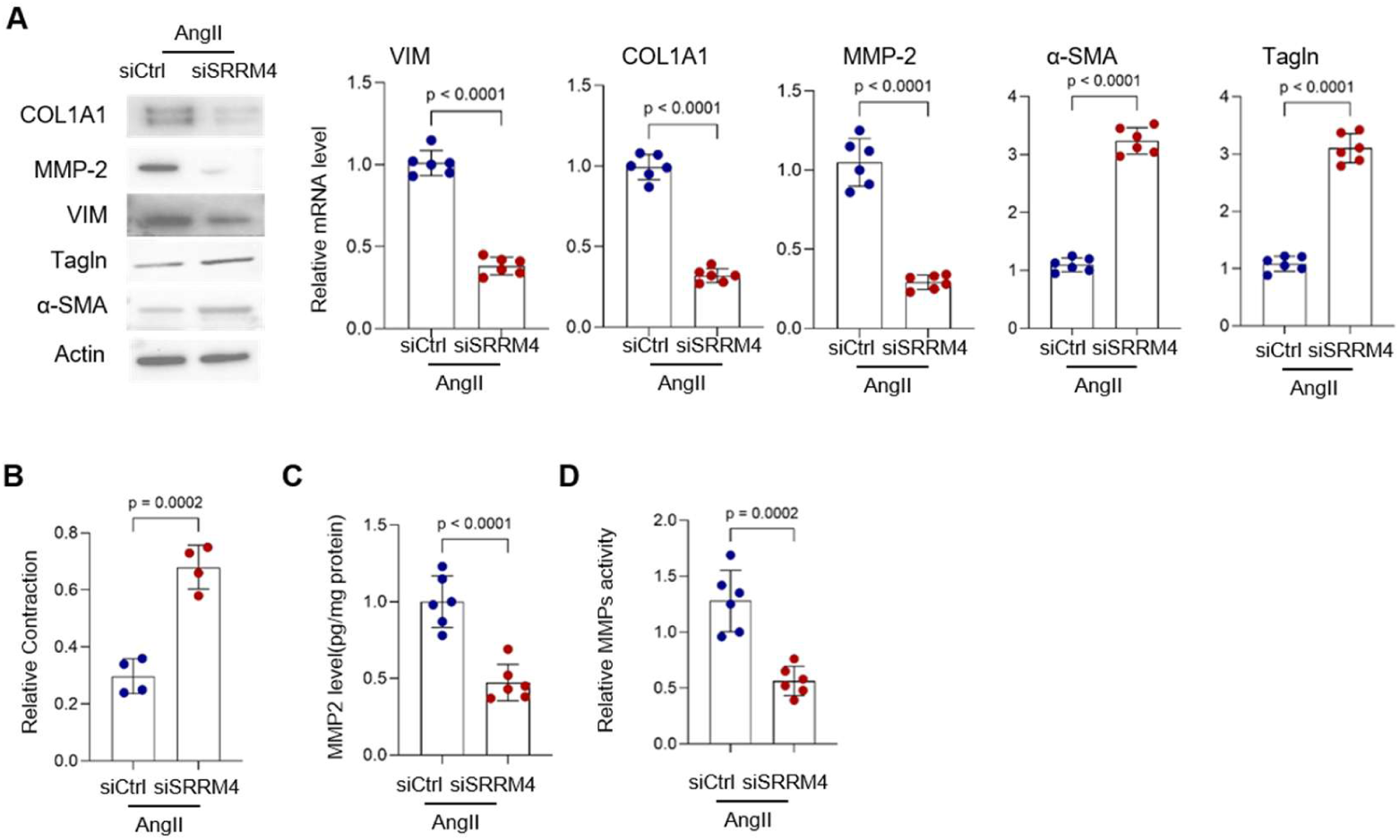
**A-D)** VSMC were stably expressed control shRNA or shRNA targeting SRRM4 and then treated with AngII for 48 hours. A) Quantification of mRNA and protein levels of α-SMA, VIM, and MMP2 in VSMC and VSMC with SRRM4 knock down; B) VSMC contraction ability were measured by collagen gel contraction assay; C) Secreted MMP2 levels were measured by ELISA; D) MMP activity of indicated cells were measured by MMP activity assays.

**Figure S15.**
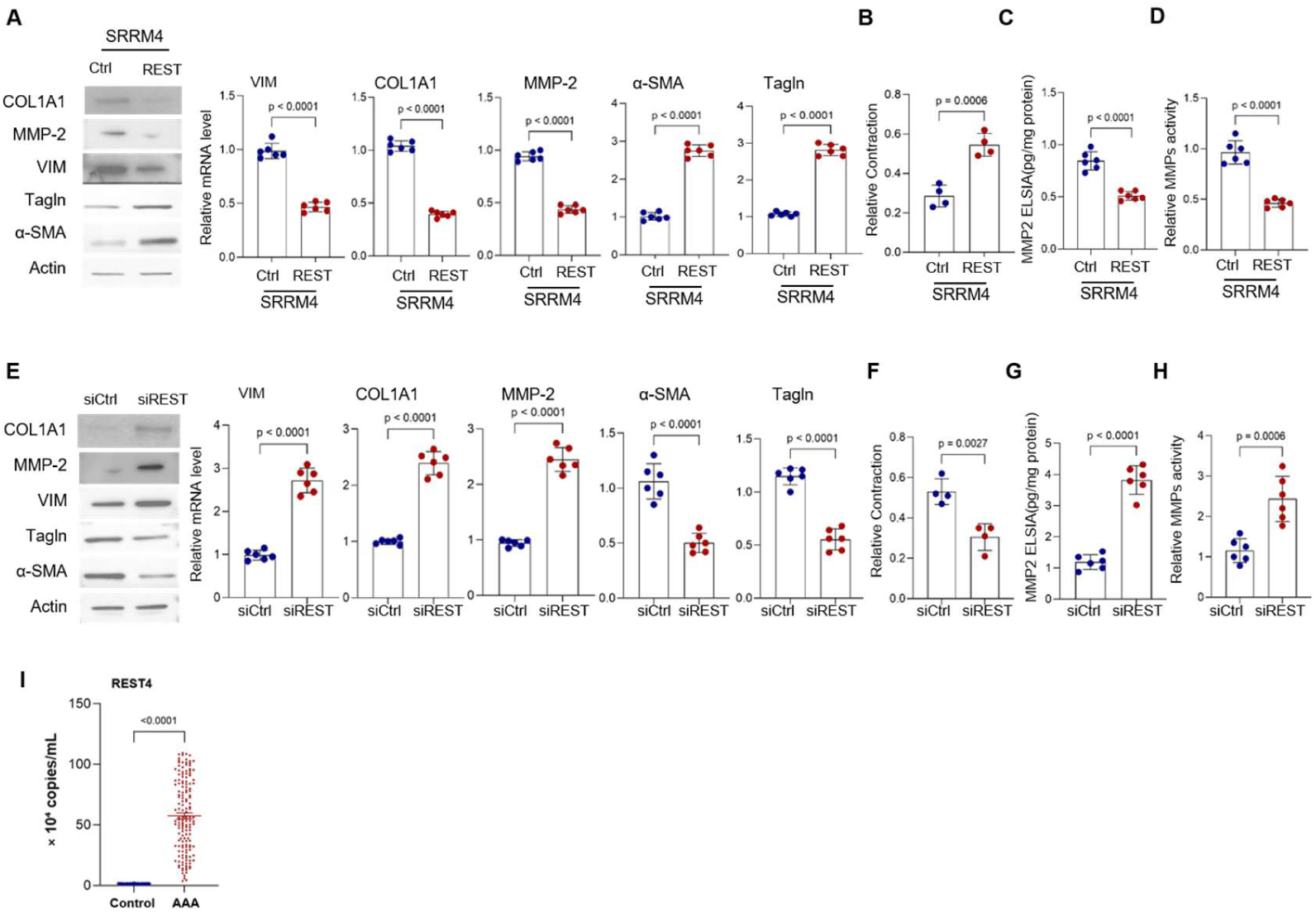
**A-D)** VSMCs with SRRM4 overexpression were transfected with either control or REST expression vectors. Quantification of A) protein and mRNA levels of α-SMA and VIM were measured by immunoblotting and realtime-qPCR, B) VSMC contraction ability were measured by collagen gel contraction assay, C) Secreted MMP2 level were measured by ELISA, D) MMP activity of indicated cells were measured by MMP activity assays. **E-H)** VSMCs were transfected with either control or siRNA targeting REST. Quantification of E) protein and mRNA levels of α-SMA and VIM were measured by immunoblotting and realtime-qPCR, F) VSMC contraction ability were measured by collagen gel contraction assay, G) Secreted MMP2 levels were measured by ELISA, H) MMP activity of indicated cells were measured by MMP activity assays. I) Plasma exosomes were isolated from 154 AAA patients and 234 age- and sex-matched healthy controls. Relative REST4 mRNA levels in exosomes were quantified by RT-qPCR.

**Figure S16.**
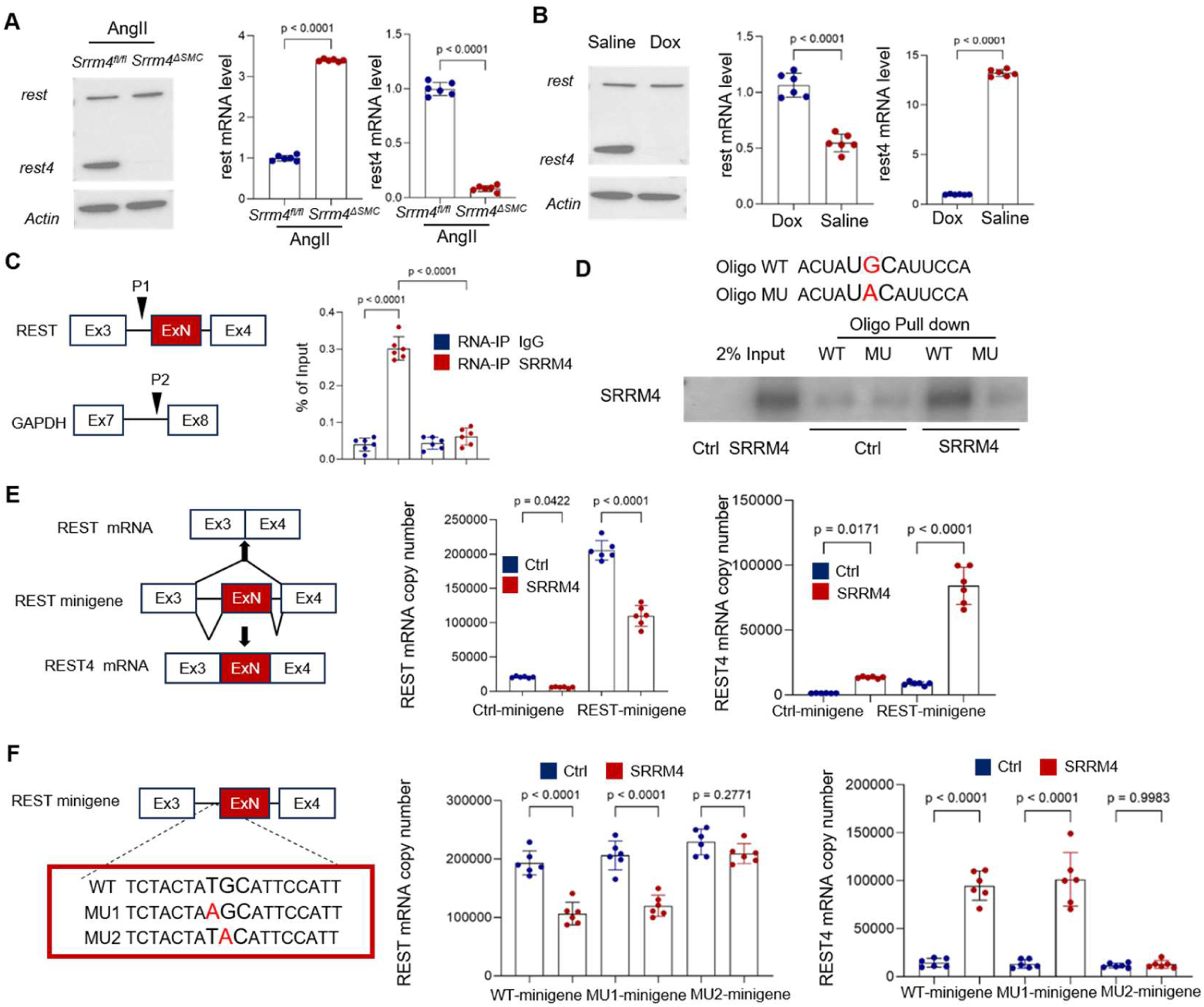
**A)** Primary cultured VSMCs derived from Saline-or AngII-treated *ApoE*^−/−^/*Srrm4^flox/flox^* and *ApoE*^−/−^/*Srrm4^ΔSMC^* mice were collected. Expression of *Rest* and *Rest4* were detected by realtime-qPCR and immunoblotting. **B)** Primary cultured VSMCs derived from *Tagln-rtTA; TRE3G-shRNA(Srrm4)* mice were treated with Saline of DOX for 7 days. Expression of *Rest* and *Rest4* were detected by realtime-qPCR and immunoblotting. **C)** A schematic diagram shows the regions (P1, P2) that were amplified in RNA-IP assays. VSMCs were transfected with Flag-SRRM4 plasmids. RNA-IP assays were performed using Flag antibody or IgG. Eluted RNA fragments were used as templates for real-time qPCR. Signals were calculated as percentage of input. **D)** Flag-tagged SRRM4 was purified from VSMCs to perform RNA pulldown assays. Oligo associated proteins were detected by immunoblotting. **E)** A schematic diagram shows the REST minigene structure and the splicing variants derived from the minigene. VSMCs were transfected with control or REST minigene reporter in the presence of -/+SRRM4. Total RNA was extracted to measure REST and REST4 mRNA levels by real-time qPCR. **F)** Site-directed mutagenesis was performed around the UGC motif in the REST minigene. VSMC were transfected with control, REST minigene, or REST minigene with indicated mutations in the presence of -/+SRRM4.Total RNA was collected and used to measure REST and REST4 mRNA levels by real-time qPCR.

**Figure S17.**
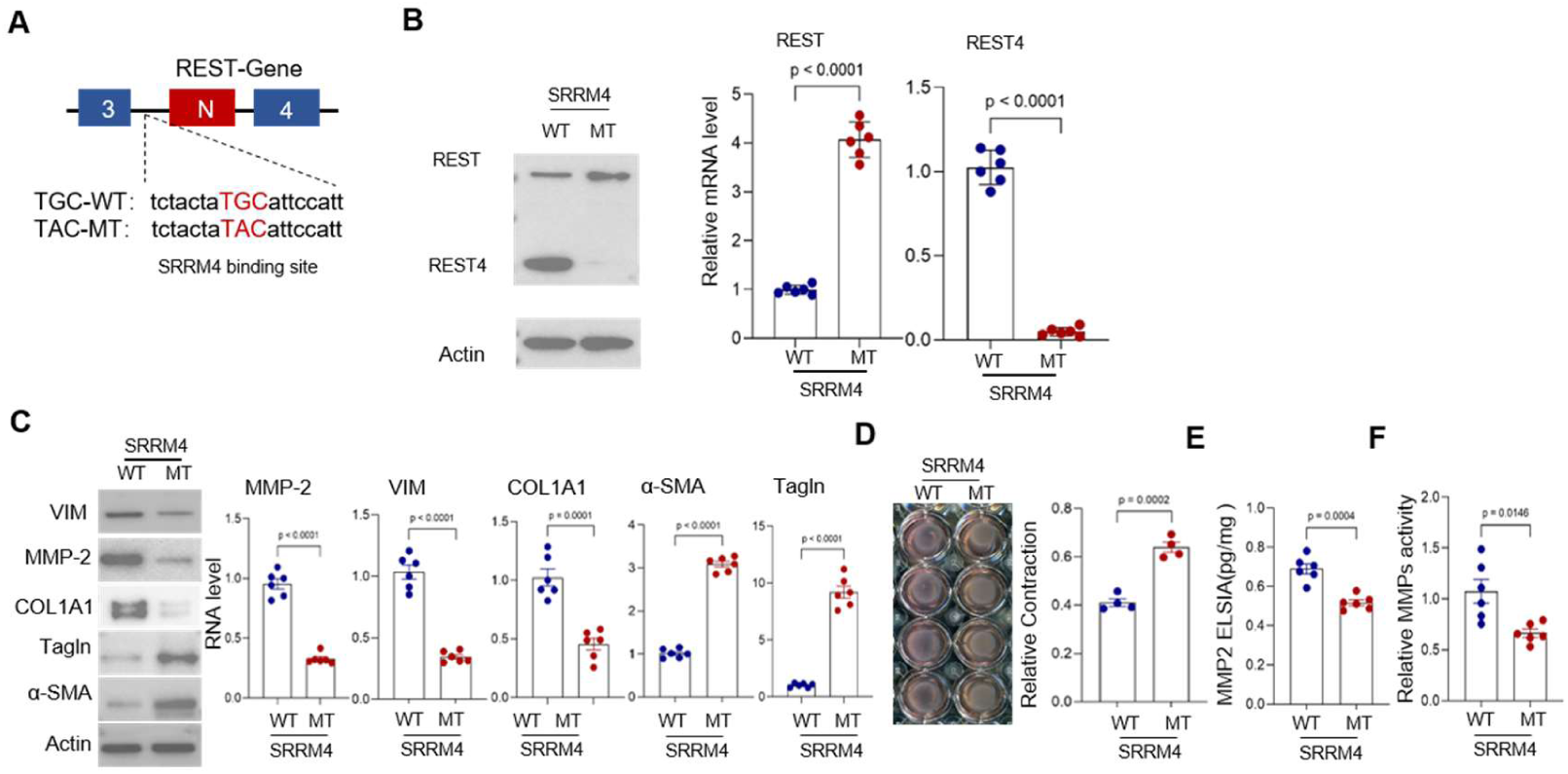
**A)** Schematic diagram of: Mutations at the TGC site in the intronic region of exon N of the REST gene in smooth muscle cells were constructed as described in the Material and Methods. **B)** Wild-type (WT) and mutant (MT) VSMCs were transfected with expression vector encoding SRRM4 protein. Expression of mRNA and protein levels of REST and REST4 were evaluated by immunoblotting and realtime-qPCR. **C-F)** VSMC(WT) and VSMC(MT) were transfected with expression vector encoding SRRM4 protein. C) Expression of mRNA and protein levels of α-SMA, Tagln, VIM, COL1A1 and MMP2 were evaluated by immunoblotting and realtime-qPCR; D) VSMC contraction ability was measured by collagen gel contraction assay; E) Secreted MMP2 level was measured by ELISA; F) MMP activity of indicated cells was measured by MMP activity assays.

**Figure S18.**
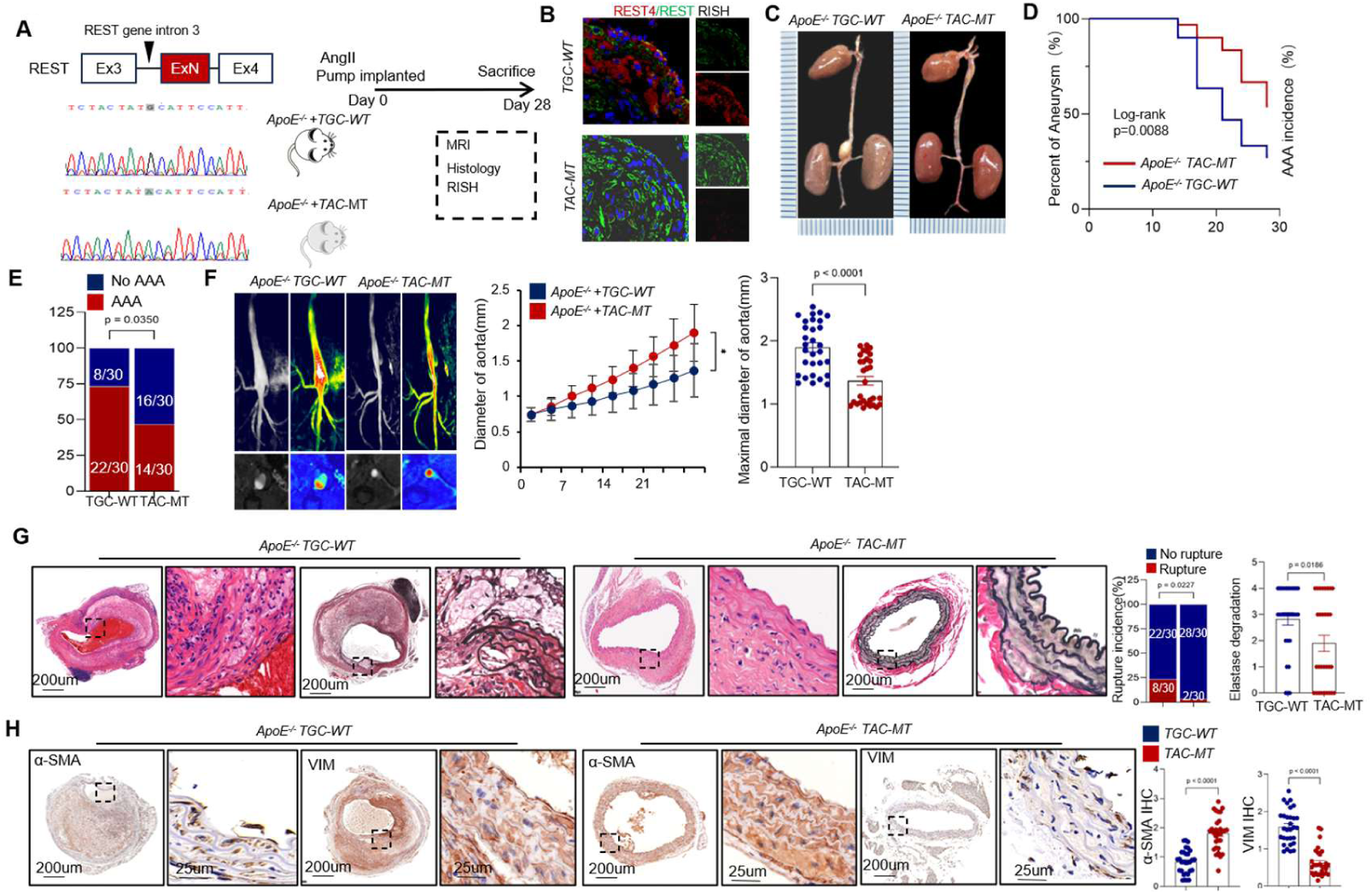
**A)** The genomic engineered mice with mutations at the TGC site in the intronic region of exon N of the REST gene in smooth muscle cells were constructed as described in the Material and Methods. Wild-type (TGC-WT) and mutant (TAC-MT) *ApoE^-/-^*mice were implanted subcutaneously with AngII pumps, and after 28 days, aortic tissue samples were collected. **B)** RISH was used to verify the specific expression of the *Rest* and *Rest4* mRNA in smooth muscle cells. **C)** Representative images of the macroscopic features of AAA formation in indicated groups. **D)** Survival curves in indicated groups. Survival data were analyzed by the Kaplan-Meier method and compared using log-rank tests. **E)** The incidence of AngII-induced aneurysm in indicated groups were analyzed by the Fisher exact test. **F)** Representative images of abdominal aortas visualized by MRI in indicated groups. Growth curve of diameter of suprarenal abdominal aortas measured by MRI in indicated groups. Quantification of the maximal diameter of suprarenal abdominal aortas from indicated groups. **G)** Representative H&E and EVG staining images of suprarenal abdominal aortas in indicated groups after 28 days of saline or AngII infusion. Quantification of elastin degradation by counting the number of breaks per vessel in suprarenal aortic segments, based on EVG staining. The rupture incidence of AngII-induced aneurysm in indicated groups was analyzed by the Fisher exact test. **H)** Representative images of *Vim* and *α-Sma* protein expression by IHC staining of indicated groups.

**Figure S19.**
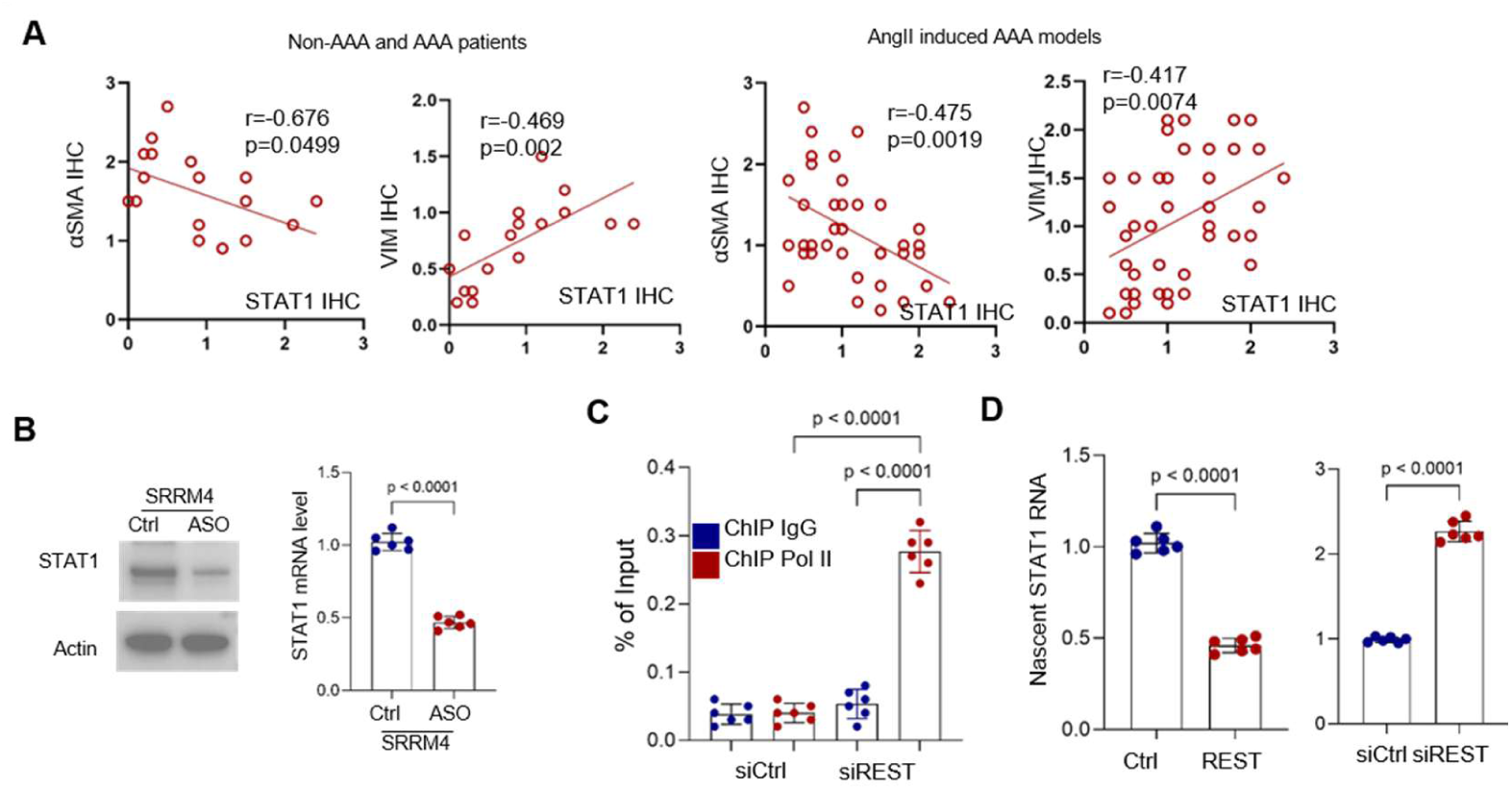
**A)** Quantification of STAT1 expression were measured by IHC in human AAA and non-AAA segments as well as abdominal aortic segments derived from *ApoE^−/−^* mice treated with saline or AngII, Pearson correlation of α-SMA and VIM with STAT1 IHC score were calculated and plotted. **B**)VSMCs stably overexpressed SRRM4 were treated with either control ASO or ASO targeting REST splicing Quantification of mRNA and protein levels of STAT1 were performed immunoblotting and realtime-qPCR. **C)** VSMCs transfected with either control siRNA or siRNA targeting REST were used to perform ChIP assays with the RNA polymerase II (Pol II) or IgG antibody. Eluted DNA fragments were used as templates for real-time qPCR to measure the enrichment of Pol II to transcription start sites (TSS) in the STAT1 gene. Signals were calculated as percentage of input. **D)** VSMCs with gain-or lost-function of REST were used for RNA run-on assay. Nascent STAT1 mRNAs were measured by real-time qPCR.

**Figure S20.**
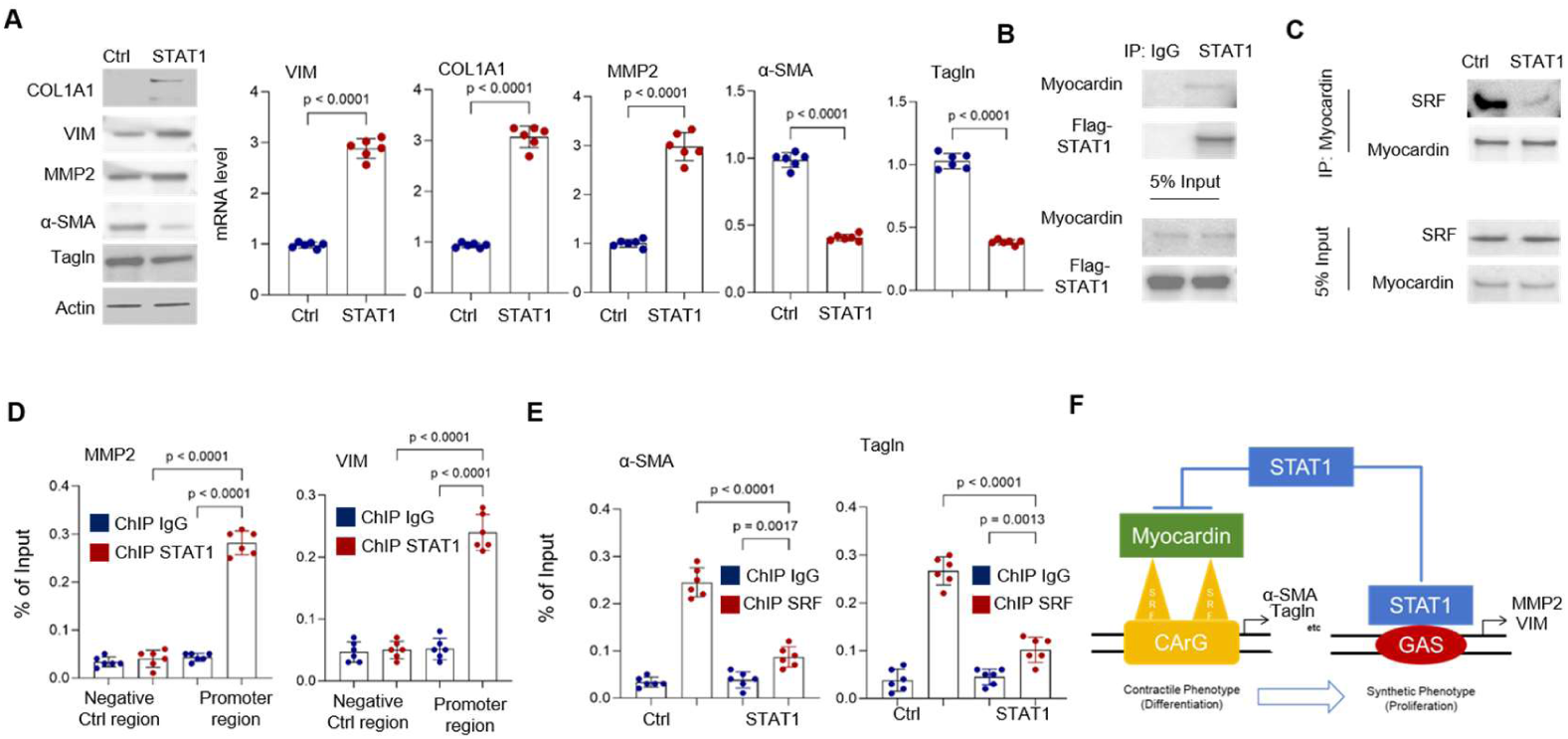
**A)** VSMCs transfected with either control vector or expressing vector encoding STAT1 gene. Expression of α-SMA, VIM, and MMP2 were detected by realtime-qPCR and immunoblotting. **B)** VSMCs with STAT1 overexpression were used to perform co-IP assay using IgG or STAT1 antibody. STAT1-associated proteins were enriched for immunoblotting assays using Myocardin antibody. **C)** VSMCs with STAT1 overexpression were used to perform co-IP assay using Myocardin antibody. Myocardin-associated proteins were enriched for immunoblotting assays using SRF antibody. **D)** VSMCs were used to perform ChIP assays with the STAT1 or IgG antibody. Eluted DNA fragments were used as templates for real-time qPCR to measure the enrichment of STAT1 to elements in the MMP2 and VIM promoter. Signals were calculated as percentage of input. **E)** VSMCs were used to perform ChIP assays with the SRF or IgG antibody. Eluted DNA fragments were used as templates for real-time qPCR to measure the enrichment of STAT1 to elements in the α-SMA and Tagln promoter. Signals were calculated as percentage of input. **F)** Diagram showed the mechanism of STAT1 regulating VSMC phenotype switch.

**Figure S21.**
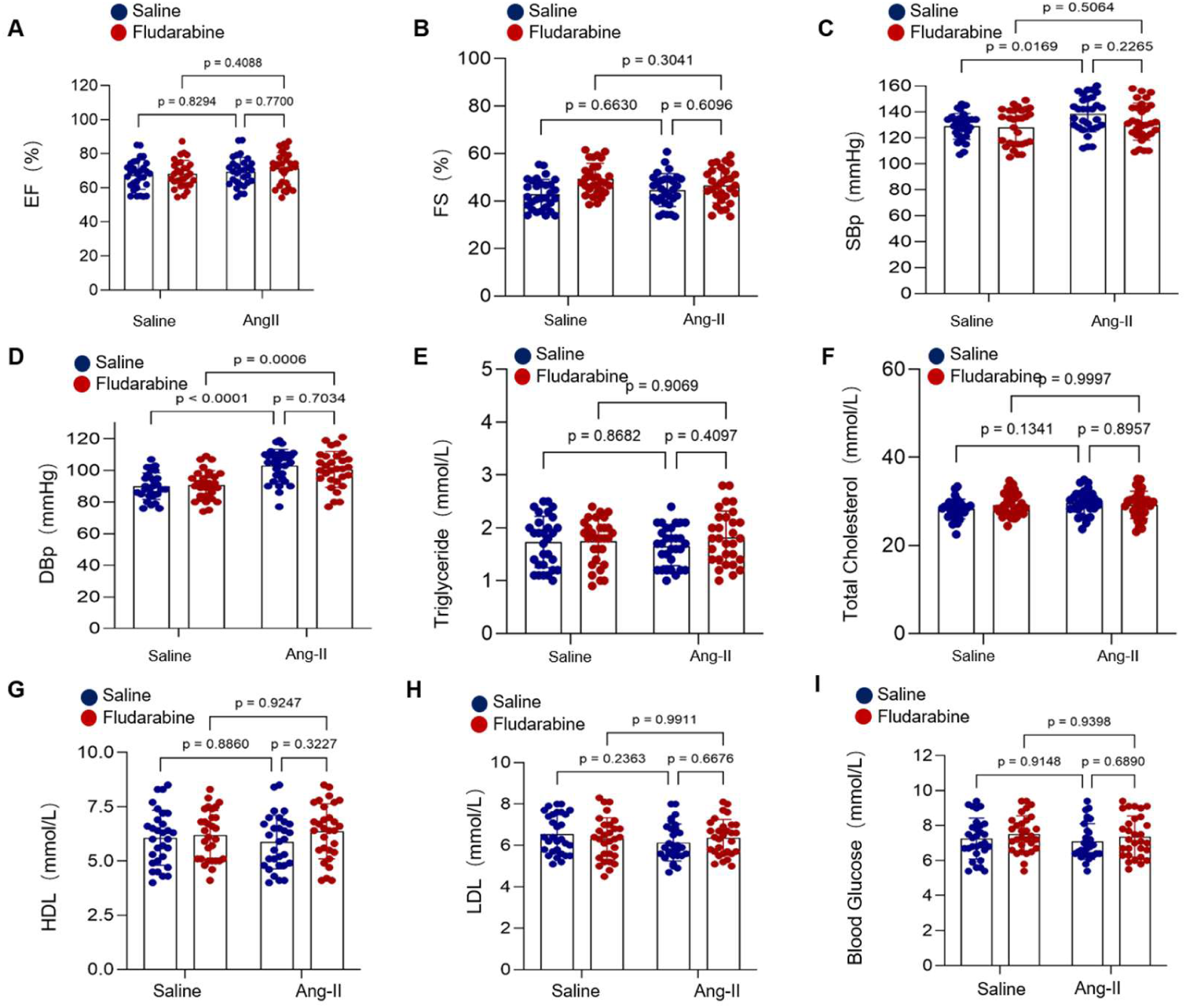
STAT1 inhibition using fludarabine 50ug/g body weight for 28 days did not overtly affect cardiac function, blood pressure, and metabolic parameters of *ApoE^−/−^* mice.

**Figure S22.**
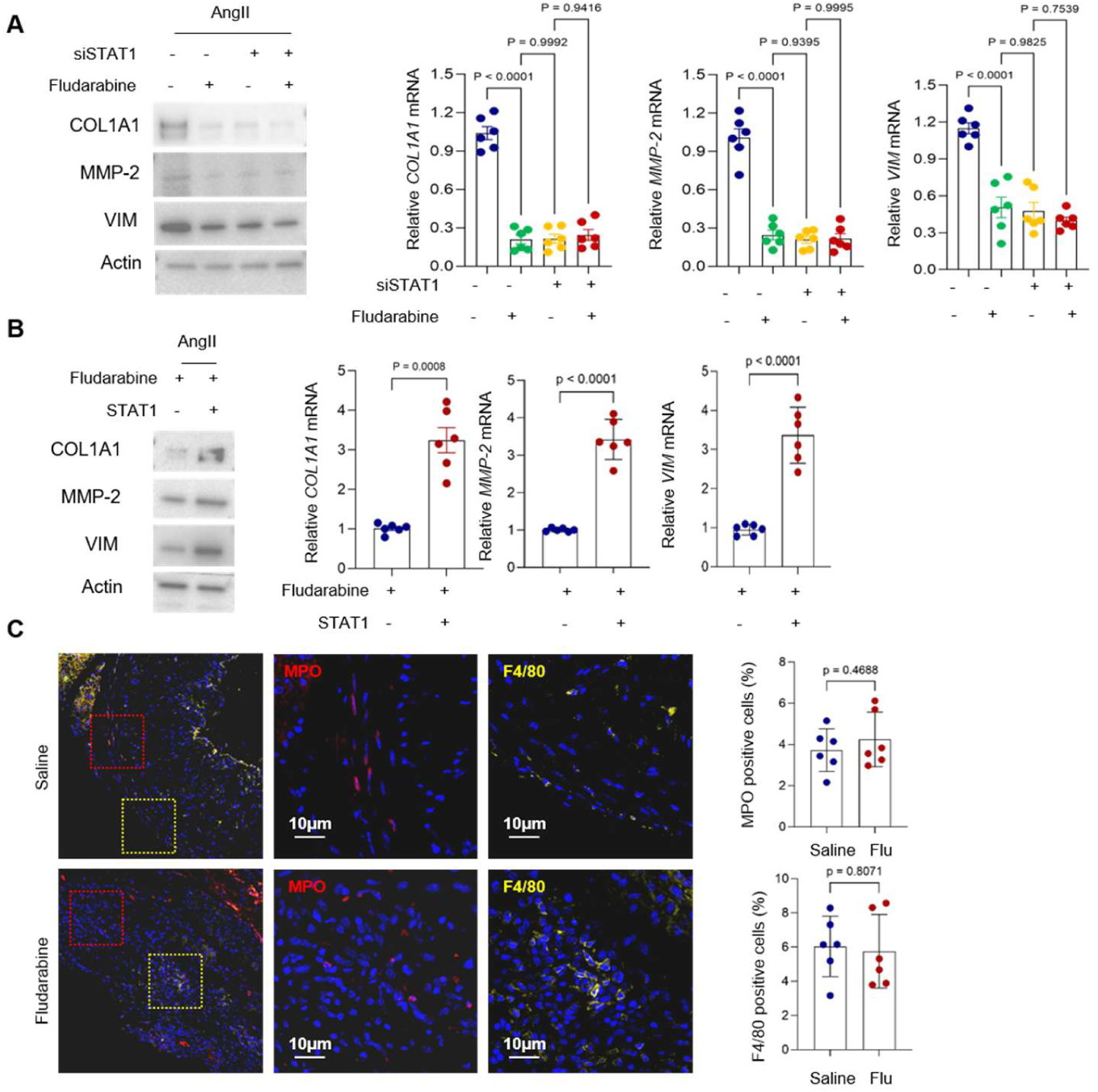
**A)** AngII-stimulated VSMCs were treated with siSTAT1, fludarabine, or siSTAT1 plus fludarabine, respectively. Expression of the synthetic markers COL1A1, MMP2, and VIM was assessed by immunoblotting and real-time PCR. **B)** Fludarabine treated VSMCs were transfected with expression vector encoding either control or STAT1, Expression of the synthetic markers COL1A1, MMP2, and VIM was assessed by immunoblotting and real-time PCR. **C)**Representative images of MPO and F4/80 expression by IF staining of *ApoE^−/−^* mice treated with vehicle or Fludarabine.

